# Extracellular Vesicles for Alzheimer’s Disease and Dementia Diagnosis

**DOI:** 10.1101/2024.04.24.24306155

**Authors:** Hash Brown Taha

**Author notes:** ORCID-ID: 0009-0007-3056-8878.

## Abstract

Accurate differential diagnosis of dementia disorders including Alzheimer’s disease (AD), frontotemporal dementia (FTD), dementia with Lewy bodies (DLB), Parkinson’s disease dementia (PDD), and vascular cognitive impairment and dementia (VCID), along with conditions like prodromal mild cognitive impairment (MCI) or negative controls (NCs), continues to challenge neurologists. The nuanced and sometimes shared pathophysiological features underscore the need for precision in developing disease-modifying therapies. In the pursuit of reliable antemortem biomarkers, extracellular vesicles (EVs) have emerged as a popular tool for their capacity to encapsulate disease-specific signatures, particularly in neurodegenerative and neurological disorders. To this end, we have performed a comprehensive, PRISMA-guided systematic review and meta-analysis, utilizing sophisticated statistical modeling to determine the diagnostic accuracy, explore between-study variance and heterogeneity (I^2^), and investigate potential publication bias using various statistical tests.

Biomarkers derived from general EVs demonstrated superior diagnostic accuracy, less between-study variance, heterogeneity, and publication bias than those from speculative CNS-enriched EVs. The trim-and-fill method suggested a potential overestimation of diagnostic effectiveness for biomarkers derived from CNS-enriched EVs due to four hypothesized missing studies with low diagnostic results, but none for general EVs. Meta-regressions revealed that studies using cerebrospinal fluid or serum, involving non-fasting individuals, sampling in the afternoon, employing citrate instead of EDTA for blood collection, using thrombin for coagulation factor depletion, isolating EVs with purer methods such as combined ultracentrifugation and size-exclusion chromatography, not freezing EVs post-isolation, and quantifying miRNA biomarkers, achieved better diagnostic accuracy and less heterogeneity. The diagnostic accuracy was low in differentiating among different dementia disorders. However, the analysis for diagnosing persons with AD vs. VCID achieved the highest diagnostic accuracy, suggesting that further studies may focus on this comparison. Additionally, we highlight several limitations in the included studies and recommend the following: Implement the use of appropriate negative controls, thorough documentation of preanalytical factors, inclusion of more dementia groups beyond AD, comprehensive reporting on pharmacological treatments, consideration of racial and ethnic minority groups, adherence to ISEV guidelines, application of the A-T-N framework, detailed documentation of dementia stages, extension of studies beyond differential diagnosis, reanalysis when postmortem definitive diagnostics become available, evaluation of prodromal conversion rates, and commitment to accurate statistical modeling and data transparency. We hope that lessons learned from this comprehensive meta-analysis can be beneficial for those attempting to discover biomarkers for AD and related dementias through EVs or alternative approaches.

## INTRODUCTION

Alzheimer’s disease (AD) is a financially, physically, and psychologically debilitating multi-factorial disorder typically characterized by neuropathological accumulation of extracellular amyloid plaques and intracellular neurofibrillary tangles, predominantly composed of amyloid-β (Aβ) and tau, respectively. While AD constitutes the bulk of dementia cases, other notable disorders include prodromal mild cognitive impairment (MCI), frontotemporal dementia (FTD), dementia with Lewy body (DLB), Parkinson’s disease dementia (PDD) and vascular cognitive impairment and dementia (VCID). Each typically presents a unique underlying pathophysiology. For example, FTD can involve other proteins such as TDP-43 and FUS [1], and areas such as frontal and temporal lobes, synucleinopathies such as PDD and DLB predominantly involve α-synuclein (α-syn) [2] and to a lesser degree tau [3], while VCID involves cerebrovascular pathology across different brain regions [4].

Despite their distinct pathophysiological features, these conditions are often misdiagnosed antemortem due to their overlapping cognitive dysfunction symptoms. Definitive diagnosis is only possible through a neuropathological examination postmortem, given the current absence of accurate and reliable antemortem biomarkers [5–9]. It should also be noted that the co-occurrence of distinct neuropathological features, such as Aβ and α-synuclein deposits and vascular dysfunction, is common, and individuals are frequently diagnosed with one or more of these diseases upon postmortem examination [10–12].

This leads to several detrimental consequences. Firstly, the inability to accurately diagnose these conditions in living persons hampers the development and evaluation of potential disease-modifying therapeutics, as treatment strategies may be misdirected or ineffective against the actual underlying pathology. This lack of specificity in diagnosis also complicates the process of stratification in clinical trials, leading to less reliable outcomes and potentially obscuring beneficial effects of treatments that might be efficacious for correctly diagnosed individuals. Furthermore, misdiagnosis can cause significant emotional distress for physicians, patients, and their families, who are often left grappling with uncertainty and the emotional toll of an unpredictable disease trajectory. Therefore, finding accurate and reliable antemortem biomarkers for diagnosing persons with dementia or predicting conversion of at-risk populations is an urgent public health need, especially during the early stages.

Extracellular vesicles (EVs) are minute, bubble-like structures, encapsulated by a phospholipid bilayer that safeguards their diverse cargo of proteins, carbohydrates, lipids, and nucleic acids [13]. Unlike cells, they do not replicate. They carry the ability to traverse the blood-brain barrier to the peripheral circulation, allowing them to transport cell-state-specific signals throughout the body. Given their potential to carry materials reflective of their cells of origin, EVs have emerged as a prominent subject in biomarker discovery for neurodegenerative and neurological disorders [14–16]. In particular, EVs enriched from the central nervous system (CNS)—referred to as ‘speculative CNS-enriched EVs’—when isolated from the blood, may offer a minimally invasive diagnostic tool. Thus, biomarker discovery for neurodegenerative disorders from general EVs and/or speculative CNS-enriched EVs, in hopes of hypothetically mirroring the neuropathological conditions present within the brain, has become a popular diagnostic approach.

A few meta-analyses have been published on this topic [17–19] focusing on the levels of specific biomarkers, especially Aβ and tau in EVs. However, to date, no detailed diagnostic accuracy meta-analysis and meta-regression have been conducted that adequately accounts for the myriad of possible covariates known to substantially influence the EVs signature. Conducting such comprehensive analysis is critical not only for evaluating and comparing biomarkers derived from both general EVs and speculative CNS-enriched EVs—thereby differentiating among individuals with prodromal or clinically diagnosed dementia and from negative controls (NCs), including biomarkers beyond Aβ and tau—but also for establishing guidelines for EV isolation and biomarker quantification. These guidelines could pinpoint which preanalytical factors and methods can enhance the diagnostic accuracy of biomarkers derived from EVs.

Given these reasons, we conducted a diagnostic accuracy meta-analysis and meta-regression focusing on studies attempting to differentiate persons with prodromal (MCI) or clinically established dementia (AD, FTD, DLB, PDD or VCID) from one another or from negative controls (NC), using biomarkers derived from general EVs or speculative CNS-enriched EVs. Although the analyses center on the diagnostic accuracy of biomarkers derived from EVs for dementia, the insights gained—particularly those derived from the meta-regressions on preanalytical factors—should be broadly applicable across various fields of biomarker discovery for neurodegenerative an non-neurodegenerative disorders that utilize EVs.

## METHODS

We performed the systematic review and meta-analysis according to the guidelines outlined in the Preferred Reporting Items for Systematic Reviews and Meta-Analyses Protocols (PRISMA). We utilized anonymized data, with no collection of personal information or involvement of human subjects. Due to the complexity and detailed nature of this meta-analysis, the protocol was not registered.

### Search Strategy

We performed a thorough search for relevant studies using specific search terms related to AD and related dementias. The search was conducted in two databases, PUBMED and EMBASE, and covered studies published from the inception of the databases until Mar 22^nd^, 2024. The comprehensive search strategy can be accessed in **Table S1**.

### Terminology

Studies employing techniques to isolate EVs without further immunocapture of cell-specific EVs are termed ‘general EVs’. Studies using enrichment antibodies to immunocapture cell-specific EVs from the CNS are referred to as ‘speculative CNS-enriched EVs’ for three main reasons. First, there is no conclusive evidence confirming that these EVs originate from the CNS. Second, the antibodies used for enriching CNS EVs, particularly from neurons (e.g., L1CAM), are not exclusive to the CNS; they are also expressed on other cell types, exist in soluble forms and have been shown to not co-elute with EVs isolated using size-exclusion chromatography (SEC) [20]. Importantly, the group that initially discovered L1CAM association with EVs [21] also later found that L1CAM is cleaved from the surface of EVs [22], further obscuring their CNS origin. Lastly, the absorption and re-release of EVs by trillions of cells, along with the recycling of their cargo [23], casts doubt on whether their cargo can accurately reflect cell-state-specific messages from the CNS.

Moreover, because EVs are believed to have a biomolecular corona [24–28], we use the term biomarkers ‘derived from’ instead of biomarkers ‘in’ or ‘within’ EVs as the biomarkers measured after lysis of EVs do not necessarily have to be encased within the EVs’ phospholipid membrane.

### Eligibility criteria

Eligible studies included in the meta-analysis must have focused on assessing biomarkers derived from general EVs and/or or speculative CNS-enriched EVs isolated from a biofluid (cerebrospinal fluid (CSF), plasma, serum, saliva or urine) for AD and at least one of the following cognitive impairment or dementia disorders: MCI, FTD, PDD, DLB and/or VCID or NCs. The studies must have included a receiver operating characteristics (ROC) analysis with accompanying area under the curve (AUC), sensitivity, specificity, and sample size for each disease. If this information was not available, we contacted the authors to either obtain the missing information, for them to perform the analysis and provide the information, or provide the dataset for us to perform the analysis using a binomial logistic regression and obtain the sensitivity and specificity that maximizes Youden’s index. All studies focusing on animals, cell lines, postmortem brain tissues, or not including the specified diseases were excluded.

For any article that provided ROC analysis for discovery and validation groups, we chose the discovery, validation or both groups depending on the size and AUC. For any article where there were univariate and combined models, we selected the combined model if the AUC was higher than the singular model. For longitudinal studies or treatment interventions, we only considered the baseline assessments.

Moreover, all authors were contacted to obtain any missing information on fasting status before biofluid collection, timing of biofluid collection, anticoagulant molecule used with plasma, defibrinating treatment (e.g., thrombin or thromboplastin-D), platelet depletion, EVs’ cargo extraction method, freezing of EVs after isolation or extraction of proteins or RNAs, genetic testing for APOE and PET imaging for Aβ or tau. Additionally, the authors were contacted for missing methodological information including centrifugation speed, duration and temperature, catalog numbers for kits used, and missing demographics including age, female %, disease duration, education length and cognitive scores (e.g., MMSE and MoCA).

### Risk of bias assessment

We did not assess risk of bias using the Quality Assessment for Diagnostic Accuracy Studies (QUADAS-2) [29] because of the complex nature of the meta-analysis including contacting the authors to obtain or clarify any missing information.

### Data synthesis

The two most common models for meta-analyses of diagnostic accuracy are the hierarchical summary ROC (HSROC) model and the bivariate random effects meta-analysis model (BRMA) [30]. Both models are equivalent when no covariates are incorporated [31]. In this study, we used the BRMA model if the number of studies was > 3. If the number of studies was ≤ 3, we used a univariate fixed-effects model [32]. We further fit the data with the unstructured or structured covariance matrix based on the lowest combination of Akaike information criteria (AIC) and Bayesian information criteria (BIC). In cases of a tie, we selected the model with the lowest AIC. Importantly, in the HSROC curve, the closer the SROC line and mean point to the upper left quadrant, the higher the accuracy is.

Heterogeneity was assessed using I^2^ statistics by Zhou & Dendukuri [33]. Publication bias [34] was assessed using Begg’s rank correlation, Egger’s and Deek’s regression and the trim-and-fill method [35]. We also conducted meta-regressions using included categorical variables to investigate the potential impact of different covariates on the absolute sensitivity and specificity.

## 3. RESULTS

The systematic review encompassed 115 studies [36–150]. Detailed information for each included study in the meta-analysis is presented in **Table 1**, while studies omitted due to incomplete inclusion criteria are listed in **Table S2**. A comprehensive summary of each model’s statistics for biomarkers derived from general EVs and speculative CNS-enriched EVs for all comparative analyses is included in **Table 2**. Descriptive statistics including the biomarker(s) used, sensitivity, specificity, diagnostic odds ratio (DOR), positive and negative likelihood ratios are included in **Table 3**.

We aimed to provide as much detail as possible by sub-stratifying the data according to fasting status, timing of biofluid collection, and specific treatments such as thromboplastin-D versus thrombin (for plasma only) and platelet depletion (also for plasma only). Additionally, we considered the type of anticoagulant molecule used (for plasma only), lysis methodology, whether the EVs were frozen post-isolation or lysis, the isolation and quantification method. Due to the limited scope of studies extending beyond persons with AD and NCs, coupled with the small sample size resulting from insufficient data, meta-regressions were exclusively conducted for this comparison to evaluate the influence of preanalytical factors on the diagnostic accuracy of biomarkers derived from general EVs and/or speculative CNS-enriched EVs.

### 3.1 AD vs. NC

Analysis of biomarkers derived from general EVs (**Table 2**) irrespective of the biofluid (**Figure 2A-C**) revealed high diagnostic accuracy, sensitivity and specificity but large between-study variance and heterogeneity for sensitivity and specificity. Analysis of biomarkers derived from speculative CNS-enriched EVs (**Figure 2D-F**) revealed high diagnostic accuracy, sensitivity, and specificity, but larger variance and heterogeneity for sensitivity and specificity despite the smaller number of studies included, suggesting that biomarkers derived from general EVs may offer more reliability.

Sub analysis by biofluid (**Figure S1A-U**) revealed that the diagnostic accuracy for both general EVs and speculative CNS-enriched EVs decreased in the following order: serum > cerebrospinal (only for general EVs) > plasma > defibrinated plasma, while all methods suffered from considerable heterogeneity (**Table 2**).

Begg’s correlation (**Figure 3A**), Egger’s (**Figure 3B**) and Deek’s (**Figure 3C-D**) regression tests revealed potential publication bias for general EVs. However, close examination of the figures suggests this bias is minimal. Moreover, the trim-and-fill method did not identify any missing studies with null or low diagnostic accuracy due to publication bias for general EVs (**Figure 3E**). In contrast, while Begg’s correlation (**Figure 3F**), Egger’s (**Figure 3G**) and Deek’s regression (**Figure 3H-I**) tests did not reveal publication bias for speculative CNS-enriched EVs, the trim– and-fill method estimated 4 missing studies with low or null diagnostic accuracy due to publication bias (see white circles in **Figure 3J**), suggesting that studies with negative or null results were less likely to be published. This is in agreement with the fact that many efforts in independent replication and validation have been met with futility for many of the studies utilizing biomarkers derived from speculative CNS-enriched EVs. This is also in support of our other meta-analyses for parkinsonian disorders of which the trim-and-fill method estimated 5 missing studies out of 16 for speculative CNS-enriched EVs [151] due to publication bias, but only 2 out of 21 for general EVs [152]. Subanalysis of publication bias by medium of isolation (**Figure S2A-E, Figure S3A-J, Figure S4A-J and Figure S5A-J**) also revealed lower publication bias for general EVs vs. speculative CNS enriched EVs.

### 3.2 AD vs. NC – Meta-Regressions

We further investigated whether preanalytical factors would influence the results as described above.

#### 3.2A Fasting Status

Surprisingly, in both biomarkers derived from general EVs (**Figure S6A-C**) and speculative CNS-enriched EVs (**Figure S7A-C**), studies with non-fasting individuals achieved a higher diagnostic accuracy (**Table S3**). As fasting is expected to reduce the noise introduced to the EVs’ cargo from dietary sources and decrease release from other cells in response to food, this unexpected result hints at the possibility that factors unrelated to the EVs’ intended cargo might inadvertently contribute to inflation of the diagnostic accuracy. It is also important to note that the lack of a fasting requirement does not imply that individuals from included groups abstained from fasting. It is plausible that some may have fasted, while some did not. Variability in the findings could potentially be attributed to outliers in the study groups who consumed specific dietary foods, affecting biomarkers derived from EVs, which would be evident in the lysate following the lysis of the EVs. The absence of documented dietary intake for the included individuals underscores the necessity for future studies to identify which foods may influence the contents from EVs or their biomolecular corona, thereby affecting diagnostic accuracy. This could be used to develop guidelines for foods to avoid prior to biofluid collection, especially for those interested in biomarkers derived from EVs.

#### 3.2B Time of Collection

Acknowledging that platelets show circadian rhythmicity, peaking in the morning [153], which in part contributes to platelets having the largest number of EVs in the blood [154], and given the circadian nature of cargoes derived from EVs [155], we considered the timing of collection in our examination of diagnostic accuracy, especially since platelet depletion is commonly overlooked before EV isolation. For both general EVs (**Figure S8A-C**) and speculative CNS-enriched EVs (**Figure S9A-C**), the diagnostic accuracy was consistently higher for studies who obtained their samples in the afternoon vs. the morning (**Table S3**). We speculated that the larger presence of morning platelet EVs or circadian rhythmicity in the blood may contribute to this effect. To test these hypotheses, we further sub-stratified the analysis by defibrinated plasma and plasma as both serum and CSF are platelet-free. Indeed, the diagnostic accuracy was higher in the afternoon than in morning for general EVs (**Figure S10A-C**) and speculative CNS-enriched EVs (**Figure S11A-C**) when only accounting for studies utilizing defibrinated plasma or plasma. However, these conclusions may be limited due to the small sample size of included studies as well as different methodologies of EV isolation and biomarker quantification. We further attempted to isolate the impact of circadian rhythmicity on the EVs’ cargo by focusing on the timing of serum or CSF collection, as platelets are not expected to be present in the serum or CSF. We were unable to perform this analysis as only one included study indicated collection of CSF in the afternoon [40], while no included studies reported collection of serum in the afternoon.

#### 3.2C Anticoagulant Molecule Mixed with Plasma

In a previous review [154], we compared the usage of different anticoagulant molecules on EV isolation and biomarker analysis, and underscored the advantage of using citrate over EDTA and heparin to reduce noise introduced to the biomarker measurements from EVs, which was recently supported by a large comparative study [156]. Our comparative analysis of the diagnostic accuracy using general EVs (**Figure S12A-C**) or speculative CNS-enriched EVs (**Figure S13A-C**) demonstrated that studies using citrate did indeed achieve a higher diagnostic accuracy compared to EDTA (**Table S3**). Although we note that only one study [63] documented the usage of citrate plasma for speculative CNS-enriched EVs, limiting any definitive conclusions. Interestingly, when only comparing studies using citrate plasma, the diagnostic accuracy of general EVs surpassed that of speculative CNS-enriched EVs, suggesting that the lower diagnostic accuracy seen in differentiating persons with AD from NCs (**Figure 2**) may be attributed to the wrong choice of anticoagulant molecule (i.e., EDTA). No included study in the meta-analysis documented the usage of heparin plasma for isolation of EVs for differentiating persons with AD from NCs, precluding our ability to include it in the comparative analysis.

#### 3.2D Coagulation depletion: Thrombin vs. Thromboplastin-D

Studies attempting to deplete the coagulation factors typically either incubate plasma with thrombin or thromboplastin-D at RT for a few minutes followed by a high-speed centrifugation to precipitate the fibrinogen pellet. Importantly, the thromboplastin-D used in the included studies was derived from rabbit brain. In one study [47], the authors showed that thromboplastin-D dissolved in PBS cross-reacts with the used antibodies for phosphorylated tau at Threonine 181 (pT181-tau) and mid-region tau using ELISA, resulting in a substantially higher noise signal in comparison to thrombin. The authors also showed that thrombin produced more consistent defibrinated clots in comparison to thromboplastin-D. As such, we tested whether this could potentially influence the diagnostic accuracy of biomarkers derived from EVs. No studies documented the usage of thromboplastin-D for general EVs, as such, we limited our analysis to speculative CNS-enriched EVs. The analysis revealed that studies utilizing thromboplastin-D had higher diagnostic accuracy (**Figure S14A-C**) but also higher generalized between-study variance (σ^2^ = 2.87) and heterogeneity (I^2^ = 73.3%) in comparison to thrombin (σ^2^ = 0.06, I^2^ = 53.4%), suggesting that diagnostic accuracy may be due to noise. Furthermore, two studies using thromboplastin-D have used ELISAs for TDP43 [52] and MMP9 [54], but did not rule out their cross-reactivity with thromboplastin-D.

Lastly, because ExoQuick is incompatible with the coagulation factors found in plasma, we attempted to sub-stratify the analysis by whether studies employed ExoQuick alongside thrombin or thromboplastin-D, as opposed to those that did not. However, the paucity of studies using ExoQuick without depleting the coagulation factors precluded our ability to perform this analysis.

#### 3.2E Platelet Depletion

We further tested whether prior depletion of platelets may affect the diagnostic accuracy using studies that documented the usage of serum or specific methodologies known to deplete platelets vs. those that did not. Prior depletion of platelets appeared to have minimal effects on the diagnostic accuracy (**Table S3**) for both general EVs (**Figure S15A-C**) and speculative CNS-enriched EVs (**Figure S16A-C**).

Platelets are significantly larger, with an estimated size range of 1–5 µm [157] compared to smaller-sized EVs, such as putative exosomes ectosomes, which are typically the focus of isolation in the majority of studies using polymer-based precipitation, ultracentrifugation (UC), or SEC. Consequently, we posited that the omission of platelet depletion could be negligible for studies focused on quantifying putative smaller-sized vs. putative larger-sized EVs. To explore this hypothesis further, we attempted to sub-stratify our analysis based on studies targeting putative larger-sized EVs using lower UC speeds (10,000-20,000xg) versus those employing higher speeds (≥ 100,000xg). However, an insufficient number of studies precluded this comparison. It is also plausible that depleting platelets may not substantially affect the accuracy of diagnostic tests relative to the presence of platelet derived EVs. Unfortunately, no included studies documented methodologies to deplete platelet EVs before isolation of general EVs or speculative CNS-enriched EVs, precluding our ability to do this comparison. Notably, one study did indeed show that platelet depletion improves detection of miRNAs derived from EVs [158], but we were not able to test this comparison in our analysis.

#### 3.2F Isolation Methodology

We further compared how different isolation methodologies would impact the diagnostic accuracy of biomarkers derived from EVs. Because the majority of studies focusing on speculative CNS-enriched EVs have used a polymer-based precipitation technique (e.g., ExoQuick) before immunoprecipitation of speculative CNS-enriched EVs, we only applied this analysis to general EVs. Our analysis revealed that the diagnostic accuracy (**Figure S17A-C**) decreased in the following order combined SEC and UC > UC > membrane affinity = polymer-based precipitation > SEC > FACS, suggesting that a combination method of SEC and UC provides the best diagnostic accuracy. This also in support of the fact that although UC and SEC provide a lower quantity of EVs, they are considered to be relatively purer [28, 159, 160].

#### 3.2G Freezing of EVs

There has been considerable debate regarding the optimal storage conditions for EVs post-isolation, with numerous metrics warranting careful consideration. These include the type and pH of the buffer in which EVs are resuspended, the EVs concentration, the nature of the biofluid and the method employed for EV isolation, and the addition of protease and phosphatase inhibitors to avert enzymatic degradation or modification. Furthermore, the use of cryoprotective agents such as trehalose and dimethyl sulfoxide, the application of lyophilization, the precise management of storage temperature and duration, and rapidity of thawing temperatures are all pivotal to consider. Notably, these factors may be influenced variably by the method used for isolation and type of intended downstream applications, such as the method used for quantification of proteins, lipids, or nucleic acids. A recent systematic study evaluated eight storage strategies, and found that storing EVs at –80°C led to a time-dependent reduction in EV concentration and purity. Additionally, there was an increase in particle size, size variability, and increased occurrence of fusion phenomena. These outcomes were observed irrespective of the storage strategy employed, suggesting that, under most conditions, the storage of EVs at –80°C is detrimental to their purity [161].

This prompted us to test whether freezing of EVs post-isolation impacts the diagnostic accuracy of biomarkers derived from EVs or speculative CNS-enriched EVs. In some studies, it was difficult to ascertain whether EVs were frozen after isolation and before extraction of cargo. All authors were contacted to clarify this point. For studies where we received no response and which did not specify a method for EV isolation (such polymer-based precipitation, UC, or SEC) prior to quantification, we categorized the EVs as ‘not frozen’. In support of the notion that freezing EVs is detrimental to their purity [161], our analyses indicated that studies not subjecting their isolated EVs to freezing achieved substantially higher accuracy (**Table S3**). This trend was consistent for general EVs (**Figure S18A-C**) and speculative CNS-enriched EVs (**Figure S19A-C**).

#### 3.2H Extraction of EVs’ cargo

To further assess whether the method of EV cargo extraction could affect the diagnostic accuracy, we attempted to compare the two most popular protein extraction methods from EVs: lysis with radioimmunoprecipitation assay buffer (RIPA) and mammalian protein extraction reagent (MPER). However, only two studies employing RIPA [89, 91] from the same research group were identified, which precluded a comparative analysis. To the best of our knowledge, no study has directly compared the efficacy of RIPA vs. MPER for lysing EVs and extracting the protein cargo, nor their impact on subsequent analyses. We also aimed to compare miRNA extraction methods involving QIAzol and TRIzol. Unfortunately, the scarcity of studies using TRIzol made this comparison unfeasible.

#### 3.2I Quantification Method

In the subsequent part of our analysis, we aimed to examine how different methods for quantifying proteins and RNA affect the diagnostic accuracy (**Table S3**). Our examination of these methods from general EVs (**Figure S20A-C**) indicated that miRNA quantification techniques, specifically SYBR Green and TaqMan qPCR, were associated with the highest diagnostic accuracy. This suggests that utilizing miRNA biomarkers derived from EVs may be the best option for future studies for differentiating AD from NCs, and possibly for other comparisons and conditions. This trend was consistent when analyzing biomarkers derived from speculative CNS-enriched EVs (**Figure S21A-C**).

#### 3.2j EV subtype and antibody clonality

To isolate speculative CNS-enriched EVs, studies typically follow one of two protocols. The first protocol isolates general EVs via standard methods like polymer-based precipitation or UC, and then applies immunoprecipitation with dynabeads-coupled to enrichment antibodies. The alternative involves an initial high-speed centrifugation to remove cell debris and putative larger-sized EVs, followed by a similar immunoprecipitation step. The most popular target is neuronal EVs using L1CAM as the marker for enrichment [162]. However, the validity of L1CAM as a marker has been questioned since the protein was found to be cleaved from the surface of EVs by the same group that discovered its association with EVs [21]. Also, a subsequent study showed that anti-L1CAM antibody clone UJ127 does not elute with EVs isolated by size-exclusion chromatography [22], exists mostly in soluble forms and cross-reacts with antibodies employed for biomarker quantification. For these reasons, many groups have been exploring alternative neuronal markers, such as ATP1A3 [80], GAP43, and NLGN3 [131], GABRD and GPR162 [81] and NRXN3 [163]. Meanwhile, markers like GLAST1 are sometimes employed to enrich speculative astrocytic EVs. However, it is important to note that no study to date has proven that the EVs isolated using these markers do indeed originate from the brain.

This prompted us to compare how different markers may compare to one another in influencing the diagnostic accuracy of biomarkers derived from speculative CNS-enriched EVs. Similar to what we reported for parkinsonian disorders [151], studies employing the anti-L1CAM antibody clone 5G3 achieved higher diagnostic accuracy (**Table S3**) than anti-L1CAM antibody clone UJ127, and both were lower than studies employing anti-GLAST1 antibody for enrichment of speculative astrocytic EVs (**Figure S22A-C**). However, because only two studies utilized the anti-L1CAM clone UJ127 antibody or the anti-GLAST1 antibody, respectively, the conclusions are limited and do not definitively establish one clone or antibody over the other.

### 3.3 AD vs. MCI

Comparative analysis of biomarkers derived from general EVs (**Figure 4A-C**) and speculative CNS-enriched EVs (**Figure 4D-F**) revealed higher diagnostic accuracy for the latter with lower heterogeneity (**Table 2**), we limited our comparison of diagnostic accuracy to plasma for general EVs (**Figure S23A-C**) vs. speculative CNS-enriched EVs (**Figure S23D-F**). Similar to the above, the diagnostic accuracy was higher for speculative CNS-enriched EVs with lower heterogeneity. No publication bias was identified in all tests for general EVs (**Figure S24A-E**). In support of the publication bias seen with speculative CNS-enriched EVs for persons with AD vs. NCs, the majority of tests did not identify publication bias (**Figure S24F-I**), but the trim-and-fill method identified one missing study out of six for speculative CNS-enriched EVs (**Figure S24J**). This suggests that studies using biomarkers derived from CNS-enriched EVs with null or low diagnostic accuracy were less likely to be published.

### 3.4 AD vs. FTD

No included studies attempted to differentiate persons with AD from FTD using biomarkers derived from speculative CNS-enriched EVs. As such, we only conducted analyses for studies using biomarkers derived from general EVs. Overall, the analyses revealed low accuracy for the models utilizing combined CSF and plasma general EVs (**Figure S25A-C**), only CSF (**Figure S25D-F**) or only plasma (**Figure S25G-I**). Although some of the tests revealed the presence of publication bias (**Figure S26A-E**), the trim-and-fill method did not identify any missing studies (**Figure S26E**).

### 3.5 AD vs. DLB/PDD

No included studies attempted to differentiate persons with AD from persons with DLB/PDD using biomarkers derived from speculative CNS-enriched EVs. As such, we only conducted analyses using biomarkers derived from general EVs. Analysis of the diagnostic accuracy (**Table 2**) revealed high diagnostic accuracy (**Figure 5A-C**), which was substantially lower for studies utilizing CSF (**Figure S27A-C**) vs. plasma (**Figure S27D-F**). All tests revealed the presence of publication bias (**Figure S28A-E**) but without any identifiable missing studies (**Figure S28E**).

### 3.6 AD vs. VCID

We focused the analysis on biomarkers derived from general EVs only due to the scarcity of studies using speculative CNS-enriched EVs for this analysis. Interestingly, despite the limited number of studies evaluating biomarkers derived from general EVs for differentiating persons with AD from persons with VCID, our analysis revealed the highest diagnostic accuracy (**Figure 6A-C**) in comparison to all other analyses, with moderate heterogeneity (**Table 2**). Although many efforts have been ongoing to identify molecular and neuroimaging biomarkers [12, 164, 165], studies focusing on biomarkers derived from EV are scarce. This suggests that researchers should invest further efforts in investigating biomarkers derived from general EVs for this comparison.

### 3.7 MCI vs. NC

Overall, the analyses (**Table 2**) revealed that biomarkers derived from general EVs (**Figure 7A-C**) offer better diagnostic accuracy than speculative CNS-enriched EVs (**Figure 7D-F**).Importantly, the partial AUC, focusing on a specific range of false positive rates within the curve for speculative CNS-enriched EVs was 0.277, suggesting that biomarkers derived from speculative CNS-enriched EVs are unreliable for differentiating persons with MCI from NCs. Because the majority of studies used plasma in comparison to CSF, defibrinated plasma, and serum for speculative CNS-enriched EVs, we did not sub-stratify the analyses by medium of isolation.

Most tests did not identify publication bias either for general EVs (**Figure S29A-E**) or speculative CNS-enriched EVs (**Figure S29F-J**). However, the trim-and-fill method revealed two missing studies with null or low diagnostic accuracy for general EVs (**Figure S29E**).

As no studies attempted to differentiate persons with MCI from FTD or DLB/PDD with either biomarkers derived from general EVs or speculative CNS-enriched EVs, we were unable to conduct further analyses.

### 3.8 MCI vs. VCID

Only a few studies attempted to differentiate persons with MCI from VCID using biomarkers derived from general EVs. The diagnostic accuracy (**Figure S30A-C**) and heterogeneity were moderate (**Table 2**).

### 3.9 FTD vs. NC

Only a few studies attempted to differentiate persons with FTD from NCs using biomarkers derived from general EVs. The diagnostic accuracy was high (**Figure S31A-C**) with moderate heterogeneity (**Table 2**) and minimal publication bias (**Figure S32A-E**), with only one missing study with null or low diagnostic accuracy (**Figure S32E**).

### 3.10 FTD vs. DLB/PDD

Only a few studies attempted to differentiate persons with FTD from persons with DLB using biomarkers derived from general EVs. The diagnostic accuracy was low (**Figure S33A-C**), suggesting this approach may not be promising for FTD vs. DLB, but the conclusions remain limited due to the small number of included studies.

### 3.11 DLB/PDD vs. NC

Only a few studies attempted to differentiate persons with DLB or PDD from NCs. The diagnostic accuracy was high (**Figure S34A-C**) with high heterogeneity and no publication bias (**Figure S35A-E**). Sub analysis by media revealed that CSF general EVs (**Figure S36A-C**) had similar diagnostic accuracy to plasma general EVs (**Figure S36D-F**).

### 3.12 VCID vs. NC

Only a few studies attempted to differentiate persons with VCID from NC using biomarkers derived from general EVs. The diagnostic accuracy (**Figure S37A-C**) and heterogeneity (**Table 2**) were moderate with no publication bias (**Figure S38A-E**).

## 4. FUTURE DIRECTIONS AND LIMITATIONS

In the course of this meta-analysis, we identified several prevalent issues across the studies that warrant attention. These challenges are critical to address to improve the accuracy, reproducibility and clinical utility of studies using biomarkers derived from general EVs and/or speculative CNS-enriched EVs for differentially diagnosing prodromal or clinically established dementia, but also for other neurodegenerative disorders and beyond.

### 4.1 Inclusion of appropriate negative controls

One of the basic yet powerful tools in experimental research for proving hypotheses is the use of appropriate negative controls. Unfortunately, when it comes to biomarkers derived from EVs, this is often neglected by researchers, peer-reviewers and editors. Given the labor– and cost-intensive nature of isolating both general EVs and speculative CNS-enriched EVs, it is imperative for studies to first evaluate the biomarkers of interest directly in the biofluid. This foundational step should be established before progressing to the isolation of general EVs and then speculative CNS-enriched EVs. If the biomarker of interest cannot differentially diagnose the disease of interest (e.g., AD) with high accuracy and reproducibility without isolating EVs, studies can then spend time and effort on isolating general EVs and/or speculative CNS-enriched EVs. Moving straight to analyzing biomarkers derived from EVs, a process that is more complex, time-consuming, and resource-intensive, is not only unnecessary but is counter-intuitive to simple scientific principles.

### 4.2 Evaluation of pT217-tau as a diagnostic marker for AD

Tau, an intrinsically disordered and natively unfolded soluble protein, is predominantly expressed in the central and peripheral nervous systems, with high abundance in neuronal cells. It is also present, albeit at lower levels, in glia such as astrocytes and oligodendrocytes. It exists in six isoforms (0N3R, 0N4R, 1N3R, 1N4R, 2N3R and 2N4R), created by alternative mRNA splicing. The function and structure of tau are regulated by phosphorylation; however, in tauopathies including AD, FTD and DLB/PDD, tau proteins can become abnormally hyperphosphorylated. This hyperphosphorylation diminishes tau’s ability to bind to microtubules, leading to the formation of intracellular neurofibrillary tangles—one of the hallmarks neuropathologies of AD [166, 167].

Phosphorylated tau at Threonine 217 (pT217-tau) has emerged as a particularly promising biofluid biomarker for AD due to several compiling findings [168–176]: 1) studies indicate its high specificity for AD, distinguishing it from other dementias and neurodegenerative disorders, 2) it demonstrates remarkable accuracy in blood and CSF samples across all stages of AD, including prodromal and 3) it performs consistently across different cohorts, clinical settings, users and various immunoassays utilizing different sets of antibodies and methodologies for detection. Because of these advantages, it is crucial that studies first attempt to rule out pT217-tau as a minimally invasive and less resource-intensive biomarker for differentially diagnosing persons with AD from other dementias, or those at-risk of phenoconversion to AD. To the best of our knowledge, only one study [81] used flow-cytometry to quantify speculative CNS-enriched EVs (GABRD+ or GPR162+) pT217-tau+ EVs isolated using UC to distinguish persons AD and NCs. However, the study did not first attempt to quantify pT217-tau in the crude biofluid or even general EVs before moving to speculative CNS-enriched EVs, which we believe would have been a possible negative control.

### 4.3 Evaluation of α-synuclein using seed amplification assays for DLB and PDD

α-Synuclein, also an intrinsically disordered and natively unfolded soluble protein, is expressed in high concentrations in the central and peripheral nervous system. The function of α-synuclein is still under debate, but it is believed to play a critical role in presynaptic vesicle release [177]. Disorders where α-synuclein oligomerizes and aggregates leading to intracellular Lewy bodies and neurites are called synucleinopathies. This group includes PD, DLB, PDD and multiple system atrophy (MSA). Notably, PD, DLB and PDD also affect the tau protein to a large degree [178–181], while in MSA, tau pathology is exceptionally rare [182–184]. Importantly, synucleinopathies may present with the same triad of parkinsonism (bradykinesia, rigidity and tremor), and as with most neurodegenerative conditions, the definitive diagnosis of a synucleinopathy can only be confirmed through a neuropathological exam [2, 185–188]. As such, there is a dire need to find accurate and reliable prodromal or early stage antemortem biomarkers [189, 190].

Recently, we have conducted the largest systematic reviews and meta-analyses for biomarkers derived from EVs or speculative CNS-enriched EVs [151, 152, 191, 192] for parkinsonian disorders, revealing low to moderate diagnostic accuracy, substantial between-study variance, heterogeneity and publication bias, especially for biomarkers derived from speculative CNS-enriched EVs [151]. This suggested that this approach is unreliable for synucleinopathies and other parkinsonian tauopathies such as corticobasal syndrome and progressive supranuclear palsy.

However, seed amplification assays (SAA) using either protein misfolding cyclic amplification (PMCA) or real-time quaking-induced conversion (RT-QuIC) is becoming popular for diagnosing synucleinopathies [193–205]. Neurologists should consider this test as a part of the diagnostic work-up for persons with dementia to rule out DLB/PDD and narrow down the differential diagnosis to other dementias such as AD.

### 4.4 Documentation of pharmacological treatments, supplementations, comorbidities, and physical activity

A common shortfall in existing studies is the lack of detailed reporting on the nature and regimen of pharmacological interventions, including drug type, dosage and treatment duration, which could considerably influence the EVs’ biomarker profile. This is especially true prior to the collection of biofluids, as active pharmaceuticals in circulation are likely to interact with biomarkers derived from EVs. Adding to this, a comprehensive account of supplementation usage (e.g., Vitamin D3, multivitamin, melatonin, etc.,) and comorbid conditions is equally important since these factors can have confounding effects on biomarkers derived from EVs, especially if there is published literature indicating a certain drug/supplement or condition that could alter the EVs’ signature. Lastly, many studies report that exercise results in the release of both general exerkines and exercise-induced myokines (i.e., exerkines coming from skeletal muscle) both in the circulation and in EVs [206–209]. As such, it would be important to document the physical activity nature of included participants. Integration of these factors will help explain how confounding variables may affect biomarkers derived from EVs in dementia and other conditions.

### 4.5 Inclusion of racial and ethnic minority groups

While AD prevalence is indeed higher among White populations in the United States, Black African Americans and Hispanic/Latinx face an incidence rate that is approximately two and one and a half times higher, respectively [210]. The lack of comprehensive reporting on race, ethnicity, and socioeconomic status in studies conducted across multiple continents makes it difficult to understand if biomarkers may be beneficial for a specific racial/ethnic group, but not others. Based on countries where the included studies were conducted, we estimate that most participants are likely of White European or Asian descent. This underscores the critical need for inclusive research practices that ensure findings also apply to of Black African American and Hispanic individuals, who exhibit a higher risk of developing AD. It is plausible to speculate that a biomarker of interest may behave differently based on these, similar to other studies investigating PET neuroimaging and genetic APOE haplotype differences [211, 212].

Unfortunately, this underrepresentation has been a major public health problem across the AD field [210, 213–216], not just those utilizing EVs.

### 4.6 Reporting of detailed methodology on the EV-TRAK platform using recommendations from ISEV-associated task forces

Because many preanalytical factors are known to influence biomarkers derived from EVs, before the collection of the biofluid, guidelines on reported EV-related parameters are increasingly established by task forces like those from the International Society for Extracellular Vesicles (ISEV) for studies using CSF [217], blood (plasma, defibrinated plasma or serum) [218, 219], urine [220–222], synovial fluid and tissue-derived. ISEV’s minimal information guidelines, first published in 2014 [223] and subsequently updated in 2018 [224] and 2023 [28], offer researchers in the EV field minimal recommendations to enhance transparency, accuracy, and reproducibility. Yet, for researchers who are truly dedicated to the scientific principles of transparency and reproducibility, platforms like EV-TRACK present a robust framework for documenting methodological details.

EV-TRACK was developed by an international consortium led by Dr. An Hendrix to catalog experimental parameters in EV research. It embraces a community consensus approach, inviting researchers to contribute data from both published and unpublished experiments. This initiative aims to foster informed discussions about relevant experimental parameters, to augment the rigor and clarity of EV studies, and to chart the progression of EV research. In our meta-analysis, and despite EV-TRACK being available since 2017, we noticed that only three recently published studies (4.8%) [46, 67, 73] included an EV-TRACK ID with documented methodologies. We encourage researchers to support EV-TRACK to improve standardization of EV research by promoting systematic reporting on the biology and methodology associated with EVs. This effort will be critical in solidifying the reliability of biomarkers derived from EVs across various studies and clinical laboratory settings.

### 4.7 Usage of the A-T-N framework

The definitive diagnosis of AD is traditionally confirmed postmortem, with clinical manifestations varying widely [225]. Before death, some individuals may have significant CNS accumulation of Aβ and tau—evident through PET imaging or CSF tests—yet remain symptom-free, while others with or without these biomarkers might experience dementia or have a heightened risk of progressing to AD [226–229]. To address this diagnostic challenge, the AD field has adopted the A-T-N framework, which categorizes AD biomarkers into three groups: Amyloid plaques (A), Tau tangles (T), and Neurodegeneration or neuronal injury (N) [230]. This approach provides a more nuanced understanding of the disease by considering these distinct yet interrelated pathological features. Despite the potential of the A-T-N framework, our meta-analysis found that only a handful of studies have employed PET imaging to distinguish between persons with positive (PET+) and negative (PET-) amyloid or tau pathology. Moreover, discussions with corresponding authors of studies incorporating PET imaging suggest that differentiating persons with PET+ from PET-based on biomarkers derived from EVs has not demonstrated a high level of accuracy. This highlights a pressing need for further research utilizing the A-T-N framework to improve the precision of AD biomarkers in clinical settings.

### 4.8 Inclusion of more dementia groups beyond AD

When a person with dementia presents in a clinical setting, neurologists can readily discern that the individual’s substantial cognitive deficits signify a departure from a negative or healthy control state. Yet, the majority of studies to date focus on AD and NC, occasionally incorporating MCI but seldom including crucial groups such as FTD, DLB/PDD, and VCID. The significant limitation here is that for biomarkers to be clinically effective, they must distinguish not just between AD and NC, but also distinguish persons with dementia from one another. As such, we encourage future studies to attempt their best in including more comparative dementia groups.

### 4.9 Documentation of EOAD vs. LOAD and early-vs. mid-vs. late-stage dementia

Our comprehension of AD rooted in the study of early-onset cases, including the first described by Alois Alzheimer, has primarily centered on what is now known as Early-Onset AD (EOAD) rather than Late-Onset AD (LOAD). The distinction between EOAD and YOAD is not just nominal; they are indeed different presentations of the disease, with varying genetic, biological, and symptomatic profiles [231]. It is important to explore whether biomarkers currently associated with AD can be universally applied across different stages and underlying causes of the disease. In our meta-analysis, we noted that only one study separated the AD group into EOAD and LOAD [39], while the other majority did not report the exact disease duration or stage for persons with dementia, precluding our ability to examine such differences.

### 4.10 Extending the approach beyond the differential diagnosis

While the primary focus of many studies has been on identifying biomarkers derived from EVs for differential diagnosis, there is a dire need to assess other metrics. These include prognostic insights, tracking disease progression, monitoring treatment responses, screening for risk, stratifying participants in clinical trials, interpreting drugs pharmacokinetics and pharmacodynamics and identifying responses to environmental toxins [232–235].

### 4.11 Reanalysis using the definitive postmortem neuropathological diagnosis

The reliance on antemortem clinical diagnoses for biomarker studies in dementia is both a practical necessity and a methodological constraint. Studies typically base diagnostic of dementia on clinical assessments such as cognitive tests and questionnaires, and further disease-specific diagnostics on additional testing such as Aβ42, tau and phosphorylated tau levels in the CSF and advanced neuroimaging. However, such methods cannot irrefutably confirm the type of dementia, as clinical symptoms and presentations often overlap among dementia subtypes, while biomarkers from the CSF and neuroimaging are not definitive [225]. The gold standard for specific dementia diagnosis remains the postmortem neuropathological examination [236].

Therefore, there is a critical need for reanalysis of the biomarker diagnostic models in light of definitive postmortem findings. When the neuropathological results become available, it is essential to rerun the diagnostic models and cut-offs while adjusting for diagnostic discrepancies This reevaluation could lead to an adjustment in the perceived accuracy of biomarkers—often revealing a decrease in diagnostic precision. With many individuals from previous studies now deceased, the opportunity for postmortem verification is easy and straight-forward. Moreover, future studies should place greater emphasis on incorporating cases with confirmed neuropathological diagnoses as opposed to those with only clinical diagnosis. Though we acknowledge that finding such samples in good quality is difficult. In this case, studies should plan to follow up with another study to validate their findings in light of the postmortem diagnosis.

### 4.12 Evaluation of prodromal conversion and progression rates

MCI often serves as a prodromal condition to dementia, commonly progressing to AD [237–239]. Although several studies have included persons with MCI aiming to differentiate it from AD, NC, and to a lower extent other dementias such as FTD, DLB/PDD, and VCID, they typically measure biomarkers at only a single timepoint before conversion. This approach fails to account for the possibility that not all persons with MCI will progress to dementia. A longitudinal study design with multiple time points would be more ideal. Such a design could track changes in biomarker levels throughout the disease course, possibly revealing fluctuations that occur before and after the conversion from MCI to dementia within the same population.

Understanding these changes over time is crucial for developing reliable predictive markers and for tailoring interventions to the disease stage.

### 4.13 Call for accurate statistical modeling and transparency

While some studies have occasionally performed ROC analyses, there is a notable shortfall in the detailing of these findings. Often, the ROC curve—critical for the validity of biomarker diagnostic studies—is omitted, or there is a failure to calculate the sensitivity and specificity that maximize Youden’s index or utilizes the highest likelihood ratios obtained using Bézier curves [240]. To address these shortcomings, studies working to identify diagnostic biomarkers derived from EVs or other biofluids should perform these analyses and report the sensitivity and specificity with appropriate cut-off values or likelihood ratios. If they encounter challenges in this analysis, the authors should be transparent and share the underlying data as a supplementary file or respond to email requests. This will enable the broader research community to validate their findings or reanalyze the data using diagnostic models, in hopes of discovering new insights.

## 5. CONCLUSION

Our meta-analysis revealed that biomarkers derived from general EVs offer better diagnostic accuracy, exhibit less heterogeneity, and demonstrate substantially lower publication bias compared to speculative CNS-enriched EVs. Additionally, we outlined several guidelines for researchers in the field of EV-derived biomarkers and encouraged the incorporation of the insights and recommendations from this meta-analysis into future studies.

## Data Availability

All data produced in the present study are available upon reasonable request to the authors

## Acknowledgment

None

## REFERENCES

[1] Ghoshal N, Cairns NJ (2013) Unravelling the mysteries of frontotemporal dementia. Mo Med 110, 411–416.

[2] Poewe W, Seppi K, Tanner CM, Halliday GM, Brundin P, Volkmann J, Schrag AE, Lang AE (2017) Parkinson disease. Nat Rev Dis Primers 3, 17013. 10.1038/nrdp.2017.13.

[3] Pan L, Meng L, He M, Zhang Z (2021) Tau in the Pathophysiology of Parkinson’s Disease. J Mol Neurosci 71, 2179–2191. 10.1007/s12031-020-01776-5.

[4] Linton AE, Weekman EM, Wilcock DM (2021) Pathologic sequelae of vascular cognitive impairment and dementia sheds light on potential targets for intervention. Cereb Circ Cogn Behav 2, 100030. 10.1016/j.cccb.2021.100030.

[5] Dubois B, Villain N, Frisoni GB, Rabinovici GD, Sabbagh M, Cappa S, Bejanin A, Bombois S, Epelbaum S, Teichmann M, Habert MO, Nordberg A, Blennow K, Galasko D, Stern Y, Rowe CC, Salloway S, Schneider LS, Cummings JL, Feldman HH (2021) Clinical diagnosis of Alzheimer’s disease: recommendations of the International Working Group. Lancet Neurol 20, 484–496. 10.1016/S1474-4422(21)00066-1.

[6] McKeith IG, Boeve BF, Dickson DW, Halliday G, Taylor JP, Weintraub D, Aarsland D, Galvin J, Attems J, Ballard CG, Bayston A, Beach TG, Blanc F, Bohnen N, Bonanni L, Bras J, Brundin P, Burn D, Chen-Plotkin A, Duda JE, El-Agnaf O, Feldman H, Ferman TJ, Ffytche D, Fujishiro H, Galasko D, Goldman JG, Gomperts SN, Graff-Radford NR, Honig LS, Iranzo A, Kantarci K, Kaufer D, Kukull W, Lee VMY, Leverenz JB, Lewis S, Lippa C, Lunde A, Masellis M, Masliah E, McLean P, Mollenhauer B, Montine TJ, Moreno E, Mori E, Murray M, O’Brien JT, Orimo S, Postuma RB, Ramaswamy S, Ross OA, Salmon DP, Singleton A, Taylor A, Thomas A, Tiraboschi P, Toledo JB, Trojanowski JQ, Tsuang D, Walker Z, Yamada M, Kosaka K (2017) Diagnosis and management of dementia with Lewy bodies: Fourth consensus report of the DLB Consortium. Neurology 89, 88–100. 10.1212/WNL.0000000000004058.

[7] Biesbroek JM, Biessels GJ (2023) Diagnosing vascular cognitive impairment: Current challenges and future perspectives. Int J Stroke 18, 36–43. 10.1177/17474930211073387.

[8] Bott NT, Radke A, Stephens ML, Kramer JH (2014) Frontotemporal dementia: diagnosis, deficits and management. Neurodegener Dis Manag 4, 439–454. 10.2217/nmt.14.34.

[9] Langa KM, Levine DA (2014) The diagnosis and management of mild cognitive impairment: a clinical review. JAMA 312, 2551–2561. 10.1001/jama.2014.13806.

[10] Nichols E, Merrick R, Hay SI, Himali D, Himali JJ, Hunter S, Keage HAD, Latimer CS, Scott MR, Steinmetz JD, Walker JM, Wharton SB, Wiedner CD, Crane PK, Keene CD, Launer LJ, Matthews FE, Schneider J, Seshadri S, White L, Brayne C, Vos T (2023) The prevalence, correlation, and co-occurrence of neuropathology in old age: harmonisation of 12 measures across six community-based autopsy studies of dementia. Lancet Healthy Longev 4, e115–e125. 10.1016/S2666-7568(23)00019-3.

[11] Trejo-Lopez JA, Yachnis AT, Prokop S (2022) Neuropathology of Alzheimer’s Disease. Neurotherapeutics 19, 173–185. 10.1007/s13311-021-01146-y.

[12] Morales CD, Cotton-Samuel D, Lao PJ, Chang JF, Pyne JD, Alshikho MJ, Lippert RV, Bista K, Hale C, Edwards NC, Igwe KC, Deters K, Zimmerman ME, Brickman AM (2024) Small vessel cerebrovascular disease is associated with cognition in prospective Alzheimer’s clinical trial participants. Alzheimers Res Ther 16, 25. 10.1186/s13195-024-01395-x.

[13] van Niel G, D’Angelo G, Raposo G (2018) Shedding light on the cell biology of extracellular vesicles. Nat Rev Mol Cell Biol 19, 213–228. 10.1038/nrm.2017.125.

[14] Dutta S, Hornung S, Taha HB, Bitan G (2023) Biomarkers for parkinsonian disorders in CNS-originating EVs: promise and challenges. Acta Neuropathologica 145, 515–540. 10.1007/s00401-023-02557-1.

[15] Khadka A, Spiers JG, Cheng L, Hill AF (2023) Extracellular vesicles with diagnostic and therapeutic potential for prion diseases. Cell Tissue Res 392, 247–267. 10.1007/s00441-022-03621-0.

[16] Hill AF (2019) Extracellular vesicles and neurodegenerative diseases. Journal of Neuroscience 39, 9269–9273. 10.1523/JNEUROSCI.0147-18.2019.

[17] Zhang X, Liu H, Huang Y, Wang R (2023) A meta-analysis of neurogenic exosomes in the diagnosis of Alzheimer’s disease. Heliyon 9, e20604. 10.1016/j.heliyon.2023.e20604.

[18] Kim KY, Shin KY, Chang KA (2021) Brain-Derived Exosomal Proteins as Effective Biomarkers for Alzheimer’s Disease: A Systematic Review and Meta-Analysis. Biomolecules 11. 10.3390/biom11070980.

[19] Liu WL, Lin HW, Lin MR, Yu Y, Liu HH, Dai YL, Chen LW, Jia WW, He XJ, Li XL, Zhu JF, Xue XH, Tao J, Chen LD (2022) Emerging blood exosome-based biomarkers for preclinical and clinical Alzheimer’s disease: a meta-analysis and systematic review. Neural Regen Res 17, 2381–2390. 10.4103/1673-5374.335832.

[20] Norman M, Ter-Ovanesyan D, Trieu W, Lazarovits R, Kowal EJK, Lee JH, Chen-Plotkin AS, Regev A, Church GM, Walt DR (2021) L1CAM is not associated with extracellular vesicles in human cerebrospinal fluid or plasma. Nat Methods 18, 631–634. 10.1038/s41592-021-01174-8.

[21] Gutwein P, Mechtersheimer S, Riedle S, Stoeck A, Gast D, Joumaa S, Zentgraf H, Fogel M, Altevogt DP (2003) ADAM10-mediated cleavage of L1 adhesion molecule at the cell surface and in released membrane vesicles. FASEB J 17, 292–294. 10.1096/fj.02-0430fje.

[22] Gutwein P, Stoeck A, Riedle S, Gast D, Runz S, Condon TP, Marme A, Phong MC, Linderkamp O, Skorokhod A, Altevogt P (2005) Cleavage of L1 in exosomes and apoptotic membrane vesicles released from ovarian carcinoma cells. Clin Cancer Res 11, 2492–2501. 10.1158/1078-0432.CCR-04-1688.

[23] Gurung S, Perocheau D, Touramanidou L, Baruteau J (2021) The exosome journey: from biogenesis to uptake and intracellular signalling. Cell Commun Signal 19, 47. 10.1186/s12964-021-00730-1.

[24] Wolf M, Poupardin RW, Ebner-Peking P, Andrade AC, Blochl C, Obermayer A, Gomes FG, Vari B, Maeding N, Eminger E, Binder HM, Raninger AM, Hochmann S, Brachtl G, Spittler A, Heuser T, Ofir R, Huber CG, Aberman Z, Schallmoser K, Volk HD, Strunk D (2022) A functional corona around extracellular vesicles enhances angiogenesis, skin regeneration and immunomodulation. J Extracell Vesicles 11, e12207. 10.1002/jev2.12207.

[25] Toth EA, Turiak L, Visnovitz T, Cserep C, Mazlo A, Sodar BW, Forsonits AI, Petovari G, Sebestyen A, Komlosi Z, Drahos L, Kittel A, Nagy G, Bacsi A, Denes A, Gho YS, Szabo-Taylor KE, Buzas EI (2021) Formation of a protein corona on the surface of extracellular vesicles in blood plasma. J Extracell Vesicles 10, e12140. 10.1002/jev2.12140.

[26] Roux Q, Boiy R, De Vuyst F, Tkach M, Pinheiro C, de Geyter S, Miinalainen I, Thery C, De Wever O, Hendrix A (2023) Depletion of soluble cytokines unlocks the immunomodulatory bioactivity of extracellular vesicles. J Extracell Vesicles 12, e12339. 10.1002/jev2.12339.

[27] Buzas EI (2022) Opportunities and challenges in studying the extracellular vesicle corona. Nat Cell Biol 24, 1322–1325. 10.1038/s41556-022-00983-z.

[28] Welsh JA, Goberdhan DCI, O’Driscoll L, Buzas EI, Blenkiron C, Bussolati B, Cai H, Di Vizio D, Driedonks TAP, Erdbrugger U, Falcon-Perez JM, Fu QL, Hill AF, Lenassi M, Lim SK, Mahoney MG, Mohanty S, Moller A, Nieuwland R, Ochiya T, Sahoo S, Torrecilhas AC, Zheng L, Zijlstra A, Abuelreich S, Bagabas R, Bergese P, Bridges EM, Brucale M, Burger D, Carney RP, Cocucci E, Crescitelli R, Hanser E, Harris AL, Haughey NJ, Hendrix A, Ivanov AR, Jovanovic-Talisman T, Kruh-Garcia NA, Ku’ulei-Lyn Faustino V, Kyburz D, Lasser C, Lennon KM, Lotvall J, Maddox AL, Martens-Uzunova ES, Mizenko RR, Newman LA, Ridolfi A, Rohde E, Rojalin T, Rowland A, Saftics A, Sandau US, Saugstad JA, Shekari F, Swift S, Ter-Ovanesyan D, Tosar JP, Useckaite Z, Valle F, Varga Z, van der Pol E, van Herwijnen MJC, Wauben MHM, Wehman AM, Williams S, Zendrini A, Zimmerman AJ, Consortium M, Thery C, Witwer KW (2024) Minimal information for studies of extracellular vesicles (MISEV2023): From basic to advanced approaches. J Extracell Vesicles 13, e12404. 10.1002/jev2.12404.

[29] Whiting PF, Rutjes AW, Westwood ME, Mallett S, Deeks JJ, Reitsma JB, Leeflang MM, Sterne JA, Bossuyt PM, Group Q (2011) QUADAS-2: a revised tool for the quality assessment of diagnostic accuracy studies. Ann Intern Med 155, 529–536. 10.7326/0003-4819-155-8-201110180-00009.

[30] Rutter CM, Gatsonis CA (2001) A hierarchical regression approach to meta-analysis of diagnostic test accuracy evaluations. Stat Med 20, 2865–2884. 10.1002/sim.942.

[31] Harbord RM, Deeks JJ, Egger M, Whiting P, Sterne JA (2007) A unification of models for meta-analysis of diagnostic accuracy studies. Biostatistics 8, 239–251. 10.1093/biostatistics/kxl004.

[32] Nyaga VN, Arbyn M (2022) Metadta: a Stata command for meta-analysis and meta-regression of diagnostic test accuracy data – a tutorial. Arch Public Health 80, 95. 10.1186/s13690-021-00747-5.

[33] Zhou Y, Dendukuri N (2014) Statistics for quantifying heterogeneity in univariate and bivariate meta-analyses of binary data: the case of meta-analyses of diagnostic accuracy. Stat Med 33, 2701–2717. 10.1002/sim.6115.

[34] Lin L, Chu H (2018) Quantifying publication bias in meta-analysis. Biometrics 74, 785–794. 10.1111/biom.12817.

[35] Shi L, Lin L (2019) The trim-and-fill method for publication bias: practical guidelines and recommendations based on a large database of meta-analyses. Medicine (Baltimore) 98, e15987. 10.1097/MD.0000000000015987.

[36] Riancho J, Vazquez-Higuera JL, Pozueta A, Lage C, Kazimierczak M, Bravo M, Calero M, Gonalezalez A, Rodriguez E, Lleo A, Sanchez-Juan P (2017) MicroRNA Profile in Patients with Alzheimer’s Disease: Analysis of miR-9-5p and miR-598 in Raw and Exosome Enriched Cerebrospinal Fluid Samples. J Alzheimers Dis 57, 483–491. 10.3233/JAD-161179.

[37] Saugstad JA, Lusardi TA, Van Keuren-Jensen KR, Phillips JI, Lind B, Harrington CA, McFarland TJ, Courtright AL, Reiman RA, Yeri AS, Kalani MYS, Adelson PD, Arango J, Nolan JP, Duggan E, Messer K, Akers JC, Galasko DR, Quinn JF, Carter BS, Hochberg FH (2017) Analysis of extracellular RNA in cerebrospinal fluid. J Extracell Vesicles 6, 1317577. 10.1080/20013078.2017.1317577.

[38] Schneider R, McKeever P, Kim T, Graff C, van Swieten JC, Karydas A, Boxer A, Rosen H, Miller BL, Laforce R, Jr., Galimberti D, Masellis M, Borroni B, Zhang Z, Zinman L, Rohrer JD, Tartaglia MC, Robertson J, Genetic FTDI (2018) Downregulation of exosomal miR-204-5p and miR-632 as a biomarker for FTD: a GENFI study. J Neurol Neurosurg Psychiatry 89, 851–858. 10.1136/jnnp-2017-317492.

[39] McKeever PM, Schneider R, Taghdiri F, Weichert A, Multani N, Brown RA, Boxer AL, Karydas A, Miller B, Robertson J, Tartaglia MC (2018) MicroRNA Expression Levels Are Altered in the Cerebrospinal Fluid of Patients with Young-Onset Alzheimer’s Disease. Mol Neurobiol 55, 8826–8841. 10.1007/s12035-018-1032-x.

[40] Spitzer P, Mulzer LM, Oberstein TJ, Munoz LE, Lewczuk P, Kornhuber J, Herrmann M, Maler JM (2019) Microvesicles from cerebrospinal fluid of patients with Alzheimer’s disease display reduced concentrations of tau and APP protein. Sci Rep 9, 7089. 10.1038/s41598-019-43607-7.

[41] Muraoka S, Jedrychowski MP, Yanamandra K, Ikezu S, Gygi SP, Ikezu T (2020) Proteomic Profiling of Extracellular Vesicles Derived from Cerebrospinal Fluid of Alzheimer’s Disease Patients: A Pilot Study. Cells 9. 10.3390/cells9091959.

[42] Utz J, Berner J, Munoz LE, Oberstein TJ, Kornhuber J, Herrmann M, Maler JM, Spitzer P (2021) Cerebrospinal Fluid of Patients With Alzheimer’s Disease Contains Increased Percentages of Synaptophysin-Bearing Microvesicles. Front Aging Neurosci 13, 682115. 10.3389/fnagi.2021.682115.

[43] Tan YJ, Wong BYX, Vaidyanathan R, Sreejith S, Chia SY, Kandiah N, Ng ASL, Zeng L (2021) Altered Cerebrospinal Fluid Exosomal microRNA Levels in Young-Onset Alzheimer’s Disease and Frontotemporal Dementia. J Alzheimers Dis Rep 5, 805–813. 10.3233/ADR-210311.

[44] Longobardi A, Nicsanu R, Bellini S, Squitti R, Catania M, Tiraboschi P, Saraceno C, Ferrari C, Zanardini R, Binetti G, Di Fede G, Benussi L, Ghidoni R (2022) Cerebrospinal Fluid EV Concentration and Size Are Altered in Alzheimer’s Disease and Dementia with Lewy Bodies. Cells 11. 10.3390/cells11030462.

[45] Chatterjee M, Ozdemir S, Kunadt M, Koel-Simmelink M, Boiten W, Piepkorn L, Pham TV, Chiasserini D, Piersma SR, Knol JC, Mobius W, Mollenhauer B, van der Flier WM, Jimenez CR, Teunissen CE, Jahn O, Schneider A (2023) C1q is increased in cerebrospinal fluid-derived extracellular vesicles in Alzheimer’s disease: A multi-cohort proteomics and immuno-assay validation study. Alzheimers Dement 19, 4828–4840. 10.1002/alz.13066.

[46] Hirschberg Y, Valle-Tamayo N, Dols-Icardo O, Engelborghs S, Buelens B, Vandenbroucke RE, Vermeiren Y, Boonen K, Mertens I (2023) Proteomic comparison between non-purified cerebrospinal fluid and cerebrospinal fluid-derived extracellular vesicles from patients with Alzheimer’s, Parkinson’s and Lewy body dementia. J Extracell Vesicles 12, e12383. 10.1002/jev2.12383.

[47] Guix FX, Corbett GT, Cha DJ, Mustapic M, Liu W, Mengel D, Chen Z, Aikawa E, Young-Pearse T, Kapogiannis D, Selkoe DJ, Walsh DM (2018) Detection of Aggregation-Competent Tau in Neuron-Derived Extracellular Vesicles. Int J Mol Sci 19. 10.3390/ijms19030663.

[48] Winston CN, Goetzl EJ, Schwartz JB, Elahi FM, Rissman RA (2019) Complement protein levels in plasma astrocyte-derived exosomes are abnormal in conversion from mild cognitive impairment to Alzheimer’s disease dementia. Alzheimers Dement (Amst) 11, 61–66. 10.1016/j.dadm.2018.11.002.

[49] Kapogiannis D, Mustapic M, Shardell MD, Berkowitz ST, Diehl TC, Spangler RD, Tran J, Lazaropoulos MP, Chawla S, Gulyani S, Eitan E, An Y, Huang CW, Oh ES, Lyketsos CG, Resnick SM, Goetzl EJ, Ferrucci L (2019) Association of Extracellular Vesicle Biomarkers With Alzheimer Disease in the Baltimore Longitudinal Study of Aging. JAMA Neurol 76, 1340–1351. 10.1001/jamaneurol.2019.2462.

[50] Cha DJ, Mengel D, Mustapic M, Liu W, Selkoe DJ, Kapogiannis D, Galasko D, Rissman RA, Bennett DA, Walsh DM (2019) miR-212 and miR-132 Are Downregulated in Neurally Derived Plasma Exosomes of Alzheimer’s Patients. Front Neurosci 13, 1208. 10.3389/fnins.2019.01208.

[51] Wang D, Wang P, Bian X, Xu S, Zhou Q, Zhang Y, Ding M, Han M, Huang L, Bi J, Jia Y, Xie Z (2020) Elevated plasma levels of exosomal BACE1-AS combined with the volume and thickness of the right entorhinal cortex may serve as a biomarker for the detection of Alzheimer’s disease. Mol Med Rep 22, 227–238. 10.3892/mmr.2020.11118.

[52] Zhang N, Gu D, Meng M, Gordon ML (2020) TDP-43 Is Elevated in Plasma Neuronal-Derived Exosomes of Patients With Alzheimer’s Disease. Front Aging Neurosci 12, 166. 10.3389/fnagi.2020.00166.

[53] Nie C, Sun Y, Zhen H, Guo M, Ye J, Liu Z, Yang Y, Zhang X (2020) Differential Expression of Plasma Exo-miRNA in Neurodegenerative Diseases by Next-Generation Sequencing. Front Neurosci 14, 438. 10.3389/fnins.2020.00438.

[54] Zhao A, Li Y, Yan Y, Qiu Y, Li B, Xu W, Wang Y, Liu J, Deng Y (2020) Increased prediction value of biomarker combinations for the conversion of mild cognitive impairment to Alzheimer’s dementia. Transl Neurodegener 9, 30. 10.1186/s40035-020-00210-5.

[55] Gu D, Liu F, Meng M, Zhang L, Gordon ML, Wang Y, Cai L, Zhang N (2020) Elevated matrix metalloproteinase-9 levels in neuronal extracellular vesicles in Alzheimer’s disease. Ann Clin Transl Neurol 7, 1681–1691. 10.1002/acn3.51155.

[56] Visconte C, Golia MT, Fenoglio C, Serpente M, Gabrielli M, Arcaro M, Sorrentino F, Busnelli M, Arighi A, Fumagalli G, Rotondo E, Rossi P, Arosio B, Scarpini E, Verderio C, Galimberti D (2023) Plasma microglial-derived extracellular vesicles are increased in frail patients with Mild Cognitive Impairment and exert a neurotoxic effect. Geroscience 45, 1557–1571. 10.1007/s11357-023-00746-0.

[57] Wang T, Yao Y, Han C, Li T, Du W, Xue J, Han Y, Cai Y (2023) MCP-1 levels in astrocyte-derived exosomes are changed in preclinical stage of Alzheimer’s disease. Front Neurol 14, 1119298. 10.3389/fneur.2023.1119298.

[58] Mankhong S, Kim S, Moon S, Choi SH, Kwak HB, Park DH, Shah P, Lee PH, Yang SW, Kang JH (2023) Circulating micro-RNAs Differentially Expressed in Korean Alzheimer’s Patients With Brain Aβ Accumulation Activate Amyloidogenesis. The journals of gerontology. Series A, Biological sciences and medical sciences 78, 292–303. 10.1093/gerona/glac106.

[59] Visconte C, Fenoglio C, Serpente M, Muti P, Sacconi A, Rigoni M, Arighi A, Borracci V, Arcaro M, Arosio B, Ferri E, Golia MT, Scarpini E, Galimberti D (2023) Altered Extracellular Vesicle miRNA Profile in Prodromal Alzheimer’s Disease. Int J Mol Sci 24. 10.3390/ijms241914749.

[60] Lugli G, Cohen AM, Bennett DA, Shah RC, Fields CJ, Hernandez AG, Smalheiser NR (2015) Plasma Exosomal miRNAs in Persons with and without Alzheimer Disease: Altered Expression and Prospects for Biomarkers. PLoS One 10, e0139233. 10.1371/journal.pone.0139233.

[61] Shi M, Kovac A, Korff A, Cook TJ, Ginghina C, Bullock KM, Yang L, Stewart T, Zheng D, Aro P, Atik A, Kerr KF, Zabetian CP, Peskind ER, Hu SC, Quinn JF, Galasko DR, Montine TJ, Banks WA, Zhang J (2016) CNS tau efflux via exosomes is likely increased in Parkinson’s disease but not in Alzheimer’s disease. Alzheimer’s and Dementia 12, 1125–1131. 10.1016/j.jalz.2016.04.003.

[62] Hosseinzadeh S, Noroozian M, Mortaz E, Mousavizadeh K (2018) Plasma microparticles in Alzheimer’s disease: The role of vascular dysfunction. Metab Brain Dis 33, 293–299. 10.1007/s11011-017-0149-3.

[63] Magalhaes CA, Campos FM, Loures CMG, Fraga VG, Oliveira ACR, Chaves AC, Rocha NP, de Souza LC, Maia RD, Guimaraes HC, Cintra MTG, Mateo ECC, Bicalho MAC, das Gracas Carvalho M, Sousa LP, Caramelli P, Gomes KB (2019) Blood neuron cell-derived microparticles as potential biomarkers in Alzheimer’s disease. Clin Chem Lab Med 57, e77–e80. 10.1515/cclm-2018-0483.

[64] Lim CZJ, Zhang Y, Chen Y, Zhao H, Stephenson MC, Ho NRY, Chen Y, Chung J, Reilhac A, Loh TP, Chen CLH, Shao H (2019) Subtyping of circulating exosome-bound amyloid beta reflects brain plaque deposition. Nat Commun 10, 1144. 10.1038/s41467-019-09030-2.

[65] Fotuhi SN, Khalaj-Kondori M, Hoseinpour Feizi MA, Talebi M (2019) Long Non-coding RNA BACE1-AS May Serve as an Alzheimer’s Disease Blood-Based Biomarker. J Mol Neurosci 69, 351–359. 10.1007/s12031-019-01364-2.

[66] Gamez-Valero A, Campdelacreu J, Rene R, Beyer K, Borras FE (2019) Comprehensive proteomic profiling of plasma-derived Extracellular Vesicles from dementia with Lewy Bodies patients. Sci Rep 9, 13282. 10.1038/s41598-019-49668-y.

[67] Gamez-Valero A, Campdelacreu J, Vilas D, Ispierto L, Rene R, Alvarez R, Armengol MP, Borras FE, Beyer K (2019) Exploratory study on microRNA profiles from plasma-derived extracellular vesicles in Alzheimer’s disease and dementia with Lewy bodies. Transl Neurodegener 8, 31. 10.1186/s40035-019-0169-5.

[68] Chanteloup G, Cordonnier M, Moreno-Ramos T, Pytel V, Matias-Guiu J, Gobbo J, Cabrera-Martin MN, Gomez-Pinedo U, Garrido C, Matias-Guiu JA (2019) Exosomal HSP70 for Monitoring of Frontotemporal Dementia and Alzheimer’s Disease: Clinical and FDG-PET Correlation. J Alzheimers Dis 71, 1263–1269. 10.3233/JAD-190545.

[69] Ellegaard Nielsen J, Sofie Pedersen K, Vestergard K, Georgiana Maltesen R, Christiansen G, Lundbye-Christensen S, Moos T, Risom Kristensen S, Pedersen S (2020) Novel Blood-Derived Extracellular Vesicle-Based Biomarkers in Alzheimer’s Disease Identified by Proximity Extension Assay. Biomedicines 8. 10.3390/biomedicines8070199.

[70] Aharon A, Spector P, Ahmad RS, Horrany N, Sabbach A, Brenner B, Aharon-Peretz J (2020) Extracellular Vesicles of Alzheimer’s Disease Patients as a Biomarker for Disease Progression. Mol Neurobiol 57, 4156–4169. 10.1007/s12035-020-02013-1.

[71] Kim KM, Meng Q, Perez de Acha O, Mustapic M, Cheng A, Eren E, Kundu G, Piao Y, Munk R, Wood WH, 3rd, De S, Noh JH, Delannoy M, Cheng L, Abdelmohsen K, Kapogiannis D, Gorospe M (2020) Mitochondrial RNA in Alzheimer’s Disease Circulating Extracellular Vesicles. Front Cell Dev Biol 8, 581882. 10.3389/fcell.2020.581882.

[72] Tanaka T, Ruifen JC, Nai YH, Tan CH, Lim CZJ, Zhang Y, Stephenson MC, Hilal S, Saridin FN, Gyanwali B, Villaraza S, Robins EG, Ihara M, Scholl M, Zetterberg H, Blennow K, Ashton NJ, Shao H, Reilhac A, Chen C (2021) Head-to-head comparison of amplified plasmonic exosome Abeta42 platform and single-molecule array immunoassay in a memory clinic cohort. Eur J Neurol 28, 1479–1489. 10.1111/ene.14704.

[73] Fitz NF, Wang J, Kamboh MI, Koldamova R, Lefterov I (2021) Small nucleolar RNAs in plasma extracellular vesicles and their discriminatory power as diagnostic biomarkers of Alzheimer’s disease. Neurobiol Dis 159, 105481. 10.1016/j.nbd.2021.105481.

[74] Longobardi A, Benussi L, Nicsanu R, Bellini S, Ferrari C, Saraceno C, Zanardini R, Catania M, Di Fede G, Squitti R, Binetti G, Ghidoni R (2021) Plasma Extracellular Vesicle Size and Concentration Are Altered in Alzheimer’s Disease, Dementia With Lewy Bodies, and Frontotemporal Dementia. Front Cell Dev Biol 9, 667369. 10.3389/fcell.2021.667369.

[75] Nielsen JE, Honore B, Vestergard K, Maltesen RG, Christiansen G, Boge AU, Kristensen SR, Pedersen S (2021) Shotgun-based proteomics of extracellular vesicles in Alzheimer’s disease reveals biomarkers involved in immunological and coagulation pathways. Sci Rep 11, 18518. 10.1038/s41598-021-97969-y.

[76] Nielsen JE, Maltesen RG, Havelund JF, Faergeman NJ, Gotfredsen CH, Vestergard K, Kristensen SR, Pedersen S (2021) Characterising Alzheimer’s disease through integrative NMR– and LC-MS-based metabolomics. Metabol Open 12, 100125. 10.1016/j.metop.2021.100125.

[77] Lucien F, Benarroch EE, Mullan A, Ali F, Boeve BF, Mielke MM, Petersen RC, Kim Y, Stang C, Camerucci E, Ross OA, Wszolek ZK, Knopman D, Bower J, Singer W, Savica R (2022) Poly (ADP-Ribose) and α-synuclein extracellular vesicles in patients with Parkinson disease: A possible biomarker of disease severity. PLoS One 17, e0264446. 10.1371/journal.pone.0264446.

[78] Tian C, Stewart T, Hong Z, Guo Z, Aro P, Soltys D, Pan C, Peskind ER, Zabetian CP, Shaw LM, Galasko D, Quinn JF, Shi M, Zhang J (2022) Blood extracellular vesicles carrying synaptic function– and brain-related proteins as potential biomarkers for Alzheimer’s disease. Alzheimers Dement. 10.1002/alz.12723.

[79] Wang L, Zhen H, Sun Y, Rong S, Li B, Song Z, Liu Z, Li Z, Ding J, Yang H, Zhang X, Sun H, Nie C (2022) Plasma Exo-miRNAs Correlated with AD-Related Factors of Chinese Individuals Involved in Abeta Accumulation and Cognition Decline. Mol Neurobiol 59, 6790–6804. 10.1007/s12035-022-03012-0.

[80] You Y, Zhang Z, Sultana N, Ericsson M, Martens YA, Sun M, Kanekiyo T, Ikezu S, Shaffer SA, Ikezu T (2023) ATP1A3 as a target for isolating neuron-specific extracellular vesicles from human brain and biofluids. Sci Adv 9, eadi3647. 10.1126/sciadv.adi3647.

[81] Guo Z, Tian C, Shi Y, Song XR, Yin W, Tao QQ, Liu J, Peng GP, Wu ZY, Wang YJ, Zhang ZX, Zhang J (2024) Blood-based CNS regionally and neuronally enriched extracellular vesicles carrying pTau217 for Alzheimer’s disease diagnosis and differential diagnosis. Acta Neuropathol Commun 12, 38. 10.1186/s40478-024-01727-w.

[82] Yang TT, Liu CG, Gao SC, Zhang Y, Wang PC (2018) The Serum Exosome Derived MicroRNA-135a, –193b, and –384 Were Potential Alzheimer’s Disease Biomarkers. Biomed Environ Sci 31, 87–96. 10.3967/bes2018.011.

[83] Agliardi C, Guerini FR, Zanzottera M, Bianchi A, Nemni R, Clerici M (2019) SNAP-25 in Serum Is Carried by Exosomes of Neuronal Origin and Is a Potential Biomarker of Alzheimer’s Disease. Mol Neurobiol 56, 5792–5798. 10.1007/s12035-019-1501-x.

[84] Barbagallo C, Mostile G, Baglieri G, Giunta F, Luca A, Raciti L, Zappia M, Purrello M, Ragusa M, Nicoletti A (2020) Specific Signatures of Serum miRNAs as Potential Biomarkers to Discriminate Clinically Similar Neurodegenerative and Vascular-Related Diseases. Cell Mol Neurobiol 40, 531–546. 10.1007/s10571-019-00751-y.

[85] Soares Martins T, Magalhaes S, Rosa IM, Vogelgsang J, Wiltfang J, Delgadillo I, Catita J, da Cruz ESOAB, Nunes A, Henriques AG (2020) Potential of FTIR Spectroscopy Applied to Exosomes for Alzheimer’s Disease Discrimination: A Pilot Study. J Alzheimers Dis 74, 391–405. 10.3233/JAD-191034.

[86] Cheng L, Vella LJ, Barnham KJ, McLean C, Masters CL, Hill AF (2020) Small RNA fingerprinting of Alzheimer’s disease frontal cortex extracellular vesicles and their comparison with peripheral extracellular vesicles. J Extracell Vesicles 9, 1766822. 10.1080/20013078.2020.1766822.

[87] Nam E, Lee YB, Moon C, Chang KA (2020) Serum Tau Proteins as Potential Biomarkers for the Assessment of Alzheimer’s Disease Progression. Int J Mol Sci 21. 10.3390/ijms21145007.

[88] Dong Z, Gu H, Guo Q, Liang S, Xue J, Yao F, Liu X, Li F, Liu H, Sun L, Zhao K (2021) Profiling of Serum Exosome MiRNA Reveals the Potential of a MiRNA Panel as Diagnostic Biomarker for Alzheimer’s Disease. Mol Neurobiol 58, 3084–3094. 10.1007/s12035-021-02323-y.

[89] Soares Martins T, Marcalo R, Ferreira M, Vaz M, Silva RM, Martins Rosa I, Vogelgsang J, Wiltfang J, da Cruz ESOAB, Henriques AG (2021) Exosomal Abeta-Binding Proteins Identified by “In Silico” Analysis Represent Putative Blood-Derived Biomarker Candidates for Alzheimer s Disease. Int J Mol Sci 22. 10.3390/ijms22083933.

[90] Durur DY, Tastan B, Ugur Tufekci K, Olcum M, Uzuner H, Karakulah G, Yener G, Genc S (2022) Alteration of miRNAs in Small Neuron-Derived Extracellular Vesicles of Alzheimer’s Disease Patients and the Effect of Extracellular Vesicles on Microglial Immune Responses. J Mol Neurosci 72, 1182–1194. 10.1007/s12031-022-02012-y.

[91] Soares Martins T, Marcalo R, da Cruz ESCB, Trindade D, Catita J, Amado F, Melo T, Rosa IM, Vogelgsang J, Wiltfang J, da Cruz ESOAB, Henriques AG (2022) Novel Exosome Biomarker Candidates for Alzheimer’s Disease Unravelled Through Mass Spectrometry Analysis. Mol Neurobiol 59, 2838–2854. 10.1007/s12035-022-02762-1.

[92] Song S, Lee JU, Jeon MJ, Kim S, Sim SJ (2022) Detection of multiplex exosomal miRNAs for clinically accurate diagnosis of Alzheimer’s disease using label-free plasmonic biosensor based on DNA-Assembled advanced plasmonic architecture. Biosens Bioelectron 199, 113864. 10.1016/j.bios.2021.113864.

[93] Tasdelen E, Ozel Kizil ET, Tezcan S, Yalap E, Bingol AP, Kutlay NY (2022) Determination of miR-373 and miR-204 levels in neuronal exosomes in Alzheimer’s disease. Turk J Med Sci 52, 1458–1467. 10.55730/1300-0144.5484.

[94] Dong Z, Gu H, Guo Q, Liu X, Li F, Liu H, Sun L, Ma H, Zhao K (2022) Circulating Small Extracellular Vesicle-Derived miR-342-5p Ameliorates Beta-Amyloid Formation via Targeting Beta-site APP Cleaving Enzyme 1 in Alzheimer’s Disease. Cells 11. 10.3390/cells11233830.

[95] Li J, Ni W, Jin D, Yu Y, Xiao MM, Zhang ZY, Zhang GJ (2023) Nanosensor-Driven Detection of Neuron-Derived Exosomal Abeta(42) with Graphene Electrolyte-Gated Transistor for Alzheimer’s Disease Diagnosis. Anal Chem 95, 5719–5728. 10.1021/acs.analchem.2c05751.

[96] Song S, Lee JU, Jeon MJ, Kim S, Lee CN, Sim SJ (2023) Precise profiling of exosomal biomarkers via programmable curved plasmonic nanoarchitecture-based biosensor for clinical diagnosis of Alzheimer’s disease. Biosens Bioelectron 230, 115269. 10.1016/j.bios.2023.115269.

[97] Ryu IS, Kim DH, Ro JY, Park BG, Kim SH, Im JY, Lee JY, Yoon SJ, Kang H, Iwatsubo T, Teunissen CE, Cho HJ, Ryu JH (2023) The microRNA-485-3p concentration in salivary exosome-enriched extracellular vesicles is related to amyloid beta deposition in the brain of patients with Alzheimer’s disease. Clin Biochem 118, 110603. 10.1016/j.clinbiochem.2023.110603.

[98] Saman S, Kim W, Raya M, Visnick Y, Miro S, Saman S, Jackson B, McKee AC, Alvarez VE, Lee NC, Hall GF (2012) Exosome-associated tau is secreted in tauopathy models and is selectively phosphorylated in cerebrospinal fluid in early Alzheimer disease. J Biol Chem 287, 3842–3849. 10.1074/jbc.M111.277061.

[99] Gui Y, Liu H, Zhang L, Lv W, Hu X (2015) Altered microRNA profiles in cerebrospinal fluid exosome in Parkinson disease and Alzheimer disease. Oncotarget 6, 37043–37053. 10.18632/oncotarget.6158.

[100] Eitan E, Hutchison ER, Marosi K, Comotto J, Mustapic M, Nigam SM, Suire C, Maharana C, Jicha GA, Liu D, Machairaki V, Witwer KW, Kapogiannis D, Mattson MP (2016) Extracellular Vesicle-Associated Abeta Mediates Trans-Neuronal Bioenergetic and Ca(2+)-Handling Deficits in Alzheimer’s Disease Models. NPJ Aging Mech Dis 2, 16019-. 10.1038/npjamd.2016.19.

[101] Derkow K, Rossling R, Schipke C, Kruger C, Bauer J, Fahling M, Stroux A, Schott E, Ruprecht K, Peters O, Lehnardt S (2018) Distinct expression of the neurotoxic microRNA family let-7 in the cerebrospinal fluid of patients with Alzheimer’s disease. PLoS One 13, e0200602. 10.1371/journal.pone.0200602.

[102] Jain G, Stuendl A, Rao P, Berulava T, Pena Centeno T, Kaurani L, Burkhardt S, Delalle I, Kornhuber J, Hull M, Maier W, Peters O, Esselmann H, Schulte C, Deuschle C, Synofzik M, Wiltfang J, Mollenhauer B, Maetzler W, Schneider A, Fischer A (2019) A combined miRNA-piRNA signature to detect Alzheimer’s disease. Transl Psychiatry 9, 250. 10.1038/s41398-019-0579-2.

[103] Liu CG, Meng S, Li Y, Lu Y, Zhao Y, Wang PC (2021) MicroRNA-135a in ABCA1-labeled Exosome is a Serum Biomarker Candidate for Alzheimer’s Disease. Biomed Environ Sci 34, 19–28. 10.3967/bes2021.004.

[104] Liu CG, Zhao Y, Lu Y, Wang PC (2021) ABCA1-Labeled Exosomes in Serum Contain Higher MicroRNA-193b Levels in Alzheimer’s Disease. Biomed Res Int 2021, 5450397. 10.1155/2021/5450397.

[105] Kumar S, Reddy PH (2021) Elevated levels of MicroRNA-455-3p in the cerebrospinal fluid of Alzheimer’s patients: A potential biomarker for Alzheimer’s disease. Biochim Biophys Acta Mol Basis Dis 1867, 166052. 10.1016/j.bbadis.2020.166052.

[106] Batabyal RA, Bansal A, Cechinel LR, Authelet K, Goldberg M, Nadler E, Keene CD, Jayadev S, Domoto-Reilly K, Li G, Peskind E, Hashimoto-Torii K, Buchwald D, Freishtat RJ (2023) Adipocyte-Derived Small Extracellular Vesicles from Patients with Alzheimer Disease Carry miRNAs Predicted to Target the CREB Signaling Pathway in Neurons. Int J Mol Sci 24. 10.3390/ijms241814024.

[107] Yakabi K, Berson E, Montine KS, Bendall SC, MacCoss MJ, Poston KL, Montine TJ (2023) Human cerebrospinal fluid single exosomes in Parkinson’s and Alzheimer’s diseases. bioRxiv. 10.1101/2023.12.22.573124.

[108] Yuyama K, Sun H, Fujii R, Hemmi I, Ueda K, Igeta Y (2024) Extracellular vesicle proteome unveils cathepsin B connection to Alzheimer’s disease pathogenesis. Brain 147, 627–636. 10.1093/brain/awad361.

[109] Winston CN, Goetzl EJ, Akers JC, Carter BS, Rockenstein EM, Galasko D, Masliah E, Rissman RA (2016) Prediction of conversion from mild cognitive impairment to dementia with neuronally derived blood exosome protein profile. Alzheimers Dement (Amst) 3, 63–72. 10.1016/j.dadm.2016.04.001.

[110] Jia L, Qiu Q, Zhang H, Chu L, Du Y, Zhang J, Zhou C, Liang F, Shi S, Wang S, Qin W, Wang Q, Li F, Wang Q, Li Y, Shen L, Wei Y, Jia J (2019) Concordance between the assessment of Abeta42, T-tau, and P-T181-tau in peripheral blood neuronal-derived exosomes and cerebrospinal fluid. Alzheimers Dement 15, 1071–1080. 10.1016/j.jalz.2019.05.002.

[111] Serpente M, Fenoglio C, D’Anca M, Arcaro M, Sorrentino F, Visconte C, Arighi A, Fumagalli GG, Porretti L, Cattaneo A, Ciani M, Zanardini R, Benussi L, Ghidoni R, Scarpini E, Galimberti D (2020) MiRNA Profiling in Plasma Neural-Derived Small Extracellular Vesicles from Patients with Alzheimer’s Disease. Cells 9. 10.3390/cells9061443.

[112] Jia L, Zhu M, Kong C, Pang Y, Zhang H, Qiu Q, Wei C, Tang Y, Wang Q, Li Y, Li T, Li F, Wang Q, Li Y, Wei Y, Jia J (2021) Blood neuro-exosomal synaptic proteins predict Alzheimer’s disease at the asymptomatic stage. Alzheimers Dement 17, 49–60. 10.1002/alz.12166.

[113] Yao PJ, Eren E, Goetzl EJ, Kapogiannis D (2021) Mitochondrial Electron Transport Chain Protein Abnormalities Detected in Plasma Extracellular Vesicles in Alzheimer’s Disease. Biomedicines 9. 10.3390/biomedicines9111587.

[114] Chi H, Yao R, Sun C, Leng B, Shen T, Wang T, Zhang S, Li M, Yang Y, Sun H, Li Z, Zhang J (2022) Blood Neuroexosomal Mitochondrial Proteins Predict Alzheimer Disease in Diabetes. Diabetes 71, 1313–1323. 10.2337/db21-0969.

[115] Alvarez XA, Winston CN, Barlow JW, Sarsoza FM, Alvarez I, Aleixandre M, Linares C, Garcia-Fantini M, Kastberger B, Winter S, Rissman RA (2022) Modulation of Amyloid-beta and Tau in Alzheimer’s Disease Plasma Neuronal-Derived Extracellular Vesicles by Cerebrolysin(R) and Donepezil. J Alzheimers Dis 90, 705–717. 10.3233/JAD-220575.

[116] Li TR, Yao YX, Jiang XY, Dong QY, Yu XF, Wang T, Cai YN, Han Y (2022) beta-Amyloid in blood neuronal-derived extracellular vesicles is elevated in cognitively normal adults at risk of Alzheimer’s disease and predicts cerebral amyloidosis. Alzheimers Res Ther 14, 66. 10.1186/s13195-022-01010-x.

[117] Li Y, Meng S, Di W, Xia M, Dong L, Zhao Y, Ling S, He J, Xue X, Chen X, Liu C (2022) Amyloid-beta protein and MicroRNA-384 in NCAM-Labeled exosomes from peripheral blood are potential diagnostic markers for Alzheimer’s disease. CNS Neurosci Ther 28, 1093–1107. 10.1111/cns.13846.

[118] Boccardi V, Poli G, Cecchetti R, Bastiani P, Scamosci M, Febo M, Mazzon E, Bruscoli S, Brancorsini S, Mecocci P (2023) miRNAs and Alzheimer’s Disease: Exploring the Role of Inflammation and Vitamin E in an Old-Age Population. Nutrients 15. 10.3390/nu15030634.

[119] C AM, F MC, C MGL, V GF, de Souza LC, H CG, M TGC, M AB, M GC, L PS, Caramelli P, K BG (2019) Microparticles are related to cognitive and functional status from normal aging to dementia. J Neuroimmunol 336, 577027. 10.1016/j.jneuroim.2019.577027.

[120] Sun R, Wang H, Shi Y, Sun Z, Jiang H, Zhang J (2020) Changes in the Morphology, Number, and Pathological Protein Levels of Plasma Exosomes May Help Diagnose Alzheimer’s Disease. J Alzheimers Dis 73, 909–917. 10.3233/JAD-190497.

[121] Perrotte M, Haddad M, Le Page A, Frost EH, Fulop T, Ramassamy C (2020) Profile of pathogenic proteins in total circulating extracellular vesicles in mild cognitive impairment and during the progression of Alzheimer’s disease. Neurobiol Aging 86, 102–111. 10.1016/j.neurobiolaging.2019.10.010.

[122] Ben Khedher MR, Haddad M, Laurin D, Ramassamy C (2021) Apolipoprotein E4-driven effects on inflammatory and neurotrophic factors in peripheral extracellular vesicles from cognitively impaired, no dementia participants who converted to Alzheimer’s disease. Alzheimers Dement (N Y) 7, e12124. 10.1002/trc2.12124.

[123] Ben Khedher MR, Haddad M, Laurin D, Ramassamy C (2021) Effect of APOE epsilon4 allele on levels of apolipoproteins E, J, and D, and redox signature in circulating extracellular vesicles from cognitively impaired with no dementia participants converted to Alzheimer’s disease. Alzheimers Dement (Amst) 13, e12231. 10.1002/dad2.12231.

[124] Sproviero D, Gagliardi S, Zucca S, Arigoni M, Giannini M, Garofalo M, Olivero M, Dell’Orco M, Pansarasa O, Bernuzzi S, Avenali M, Cotta Ramusino M, Diamanti L, Minafra B, Perini G, Zangaglia R, Costa A, Ceroni M, Perrone-Bizzozero NI, Calogero RA, Cereda C (2021) Different miRNA Profiles in Plasma Derived Small and Large Extracellular Vesicles from Patients with Neurodegenerative Diseases. Int J Mol Sci 22. 10.3390/ijms22052737.

[125] Cai H, Pang Y, Wang Q, Qin W, Wei C, Li Y, Li T, Li F, Wang Q, Li Y, Wei Y, Jia L (2022) Proteomic profiling of circulating plasma exosomes reveals novel biomarkers of Alzheimer’s disease. Alzheimers Res Ther 14, 181. 10.1186/s13195-022-01133-1.

[126] Pounders J, Hill EJ, Hooper D, Zhang X, Biesiada J, Kuhnell D, Greenland HL, Esfandiari L, Timmerman E, Foster F, Wang C, Walsh KB, Shatz R, Woo D, Medvedovic M, Langevin S, Sawyer RP (2022) MicroRNA expression within neuronal-derived small extracellular vesicles in frontotemporal degeneration. Medicine (Baltimore) 101, e30854. 10.1097/MD.0000000000030854.

[127] Sproviero D, Gagliardi S, Zucca S, Arigoni M, Giannini M, Garofalo M, Fantini V, Pansarasa O, Avenali M, Ramusino MC, Diamanti L, Minafra B, Perini G, Zangaglia R, Costa A, Ceroni M, Calogero RA, Cereda C (2022) Extracellular Vesicles Derived From Plasma of Patients With Neurodegenerative Disease Have Common Transcriptomic Profiling. Front Aging Neurosci 14, 785741. 10.3389/fnagi.2022.785741.

[128] Wang Y, Yuan P, Ding L, Zhu J, Qi X, Zhang Y, Li Y, Xia X, Zheng JC (2022) Circulating extracellular vesicle-containing microRNAs reveal potential pathogenesis of Alzheimer’s disease. Front Cell Neurosci 16, 955511. 10.3389/fncel.2022.955511.

[129] Krishna G, Santhoshkumar R, Sivakumar PT, Alladi S, Mahadevan A, Dahale AB, Arshad F, Subramanian S (2023) Pathological (Dis)Similarities in Neuronal Exosome-Derived Synaptic and Organellar Marker Levels Between Alzheimer’s Disease and Frontotemporal Dementia. J Alzheimers Dis 94, S387–S397. 10.3233/JAD-220829.

[130] Kumar A, Su Y, Sharma M, Singh S, Kim S, Peavey JJ, Suerken CK, Lockhart SN, Whitlow CT, Craft S, Hughes TM, Deep G (2023) MicroRNA expression in extracellular vesicles as a novel blood-based biomarker for Alzheimer’s disease. Alzheimers Dement 19, 4952–4966. 10.1002/alz.13055.

[131] Eitan E, Thornton-Wells T, Elgart K, Erden E, Gershun E, Levine A, Volpert O, Azadeh M, Smith DG, Kapogiannis D (2023) Synaptic proteins in neuron-derived extracellular vesicles as biomarkers for Alzheimer’s disease: novel methodology and clinical proof of concept. Extracell Vesicles Circ Nucl Acids 4, 133–150. 10.20517/evcna.2023.13.

[132] Manolopoulos A, Delgado-Peraza F, Mustapic M, Pucha KA, Nogueras-Ortiz C, Daskalopoulos A, Knight DD, Leoutsakos JM, Oh ES, Lyketsos CG, Kapogiannis D (2023) Comparative assessment of Alzheimer’s disease-related biomarkers in plasma and neuron-derived extracellular vesicles: a nested case-control study. Front Mol Biosci 10, 1254834. 10.3389/fmolb.2023.1254834.

[133] Sun Y, Hefu Z, Li B, Lifang W, Zhijie S, Zhou L, Deng Y, Zhili L, Ding J, Li T, Zhang W, Chao N, Rong S (2023) Plasma Extracellular Vesicle MicroRNA Analysis of Alzheimer’s Disease Reveals Dysfunction of a Neural Correlation Network. Research (Wash D C) 6, 0114. 10.34133/research.0114.

[134] Plaschke K, Kopitz J, Gebert J, Wolf ND, Wolf RC (2023) Proteomic Analysis Reveals Potential Exosomal Biomarkers in Patients With Sporadic Alzheimer Disease. Alzheimer Dis Assoc Disord 37, 315–321. 10.1097/WAD.0000000000000589.

[135] Soares Martins T, Pelech S, Ferreira M, Pinho B, Leandro K, de Almeida LP, Breitling B, Hansen N, Esselmann H, Wiltfang J, da Cruz ESOAB, Henriques AG (2024) Phosphoproteome Microarray Analysis of Extracellular Particles as a Tool to Explore Novel Biomarker Candidates for Alzheimer’s Disease. Int J Mol Sci 25. 10.3390/ijms25031584.

[136] Phu Pham LH, Chang CF, Tuchez K, Chen Y (2023) Assess Alzheimer’s Disease via Plasma Extracellular Vesicle-derived mRNA. medRxiv. 10.1101/2023.12.26.23299985.

[137] Liu CG, Song J, Zhang YQ, Wang PC (2014) MicroRNA-193b is a regulator of amyloid precursor protein in the blood and cerebrospinal fluid derived exosomal microRNA-193b is a biomarker of Alzheimer’s disease. Mol Med Rep 10, 2395–2400. 10.3892/mmr.2014.2484.

[138] Cheng L, Doecke JD, Sharples RA, Villemagne VL, Fowler CJ, Rembach A, Martins RN, Rowe CC, Macaulay SL, Masters CL, Hill AF, Australian Imaging B, Lifestyle Research G (2015) Prognostic serum miRNA biomarkers associated with Alzheimer’s disease shows concordance with neuropsychological and neuroimaging assessment. Mol Psychiatry 20, 1188–1196. 10.1038/mp.2014.127.

[139] Wei H, Xu Y, Xu W, Zhou Q, Chen Q, Yang M, Feng F, Liu Y, Zhu X, Yu M, Li Y (2018) Serum Exosomal miR-223 Serves as a Potential Diagnostic and Prognostic Biomarker for Dementia. Neuroscience 379, 167–176. 10.1016/j.neuroscience.2018.03.016.

[140] Cicognola C, Brinkmalm G, Wahlgren J, Portelius E, Gobom J, Cullen NC, Hansson O, Parnetti L, Constantinescu R, Wildsmith K, Chen HH, Beach TG, Lashley T, Zetterberg H, Blennow K, Hoglund K (2019) Novel tau fragments in cerebrospinal fluid: relation to tangle pathology and cognitive decline in Alzheimer’s disease. Acta Neuropathol 137, 279–296. 10.1007/s00401-018-1948-2.

[141] Haddad M, Perrotte M, Landri S, Lepage A, Fulop T, Ramassamy C (2019) Circulating and Extracellular Vesicles Levels of N-(1-Carboxymethyl)-L-Lysine (CML) Differentiate Early to Moderate Alzheimer’s Disease. J Alzheimers Dis 69, 751–762. 10.3233/JAD-181272.

[142] Li F, Xie XY, Sui XF, Wang P, Chen Z, Zhang JB (2020) Profile of Pathogenic Proteins and MicroRNAs in Plasma-derived Extracellular Vesicles in Alzheimer’s Disease: A Pilot Study. Neuroscience 432, 240–246. 10.1016/j.neuroscience.2020.02.044.

[143] Arioz BI, Tufekci KU, Olcum M, Durur DY, Akarlar BA, Ozlu N, Bagriyanik HA, Keskinoglu P, Yener G, Genc S (2021) Proteome profiling of neuron-derived exosomes in Alzheimer’s disease reveals hemoglobin as a potential biomarker. Neurosci Lett 755, 135914. 10.1016/j.neulet.2021.135914.

[144] Odaka H, Hiemori K, Shimoda A, Akiyoshi K, Tateno H (2021) Platelet-derived extracellular vesicles are increased in sera of Alzheimer’s disease patients, as revealed by Tim4-based assays. FEBS Open Bio 11, 741–752. 10.1002/2211-5463.13068.

[145] Haddad M, Perrotte M, Ben Khedher MR, Madec E, Lepage A, Fulop T, Ramassamy C (2021) Levels of Receptor for Advanced Glycation End Products and Glyoxalase-1 in the Total Circulating Extracellular Vesicles from Mild Cognitive Impairment and Different Stages of Alzheimer’s Disease Patients. J Alzheimers Dis 84, 227–237. 10.3233/JAD-210441.

[146] Chen B, Song L, Yang J, Zhou WY, Cheng YY, Lai YJ (2023) Proteomics of serum exosomes identified fibulin-1 as a novel biomarker for mild cognitive impairment. Neural Regen Res 18, 587–593. 10.4103/1673-5374.347740.

[147] Zhong J, Ren X, Liu W, Wang S, Lv Y, Nie L, Lin R, Tian X, Yang X, Zhu F, Liu J (2021) Discovery of Novel Markers for Identifying Cognitive Decline Using Neuron-Derived Exosomes. Front Aging Neurosci 13, 696944. 10.3389/fnagi.2021.696944.

[148] Zhang M, Gong W, Zhang D, Ji M, Chen B, Chen B, Li X, Zhou Y, Dong C, Wen G, Zhan X, Wu X, Cui L, Feng Y, Wang S, Yuan H, Xu E, Xia M, Verkhratsky A, Li B (2022) Ageing related thyroid deficiency increases brain-targeted transport of liver-derived ApoE4-laden exosomes leading to cognitive impairment. Cell Death Dis 13, 406. 10.1038/s41419-022-04858-x.

[149] Rani K, Rastogi S, Vishwakarma P, Bharti PS, Sharma V, Renu K, Modi GP, Vishnu VY, Chatterjee P, Dey AB, Nikolajeff F, Kumar S (2021) A novel approach to correlate the salivary exosomes and their protein cargo in the progression of cognitive impairment into Alzheimer’s disease. J Neurosci Methods 347, 108980. 10.1016/j.jneumeth.2020.108980.

[150] Sun R, Wang H, Shi Y, Gao D, Sun Z, Chen Z, Jiang H, Zhang J (2019) A Pilot Study of Urinary Exosomes in Alzheimer’s Disease. Neurodegener Dis 19, 184–191. 10.1159/000505851.

[151] Taha HB, Bogoniewski A (2024) Analysis of biomarkers in speculative CNS-enriched extracellular vesicles for parkinsonian disorders: a comprehensive systematic review and diagnostic meta-analysis. J Neurol 271, 1680–1706. 10.1007/s00415-023-12093-3.

[152] Taha HB, Bogoniewski A (2023) Extracellular vesicles from bodily fluids for the accurate diagnosis of Parkinson’s disease and related disorders: A systematic review and diagnostic meta-analysis. Journal of Extracellular Biology 2, e121. 10.1002/jex2.121.

[153] Scheer FA, Michelson AD, Frelinger AL, 3rd, Evoniuk H, Kelly EE, McCarthy M, Doamekpor LA, Barnard MR, Shea SA (2011) The human endogenous circadian system causes greatest platelet activation during the biological morning independent of behaviors. PLoS One 6, e24549. 10.1371/journal.pone.0024549.

[154] Taha HB (2023) Plasma versus serum for extracellular vesicle (EV) isolation: A duel for reproducibility and accuracy for CNS-originating EVs biomarker analysis. J Neurosci Res 101, 1677–1686. 10.1002/jnr.25231.

[155] Yeung CC, Dondelinger F, Schoof EM, Georg B, Lu Y, Zheng Z, Zhang J, Hannibal J, Fahrenkrug J, Kjaer M (2022) Circadian regulation of protein cargo in extracellular vesicles. Sci Adv 8, eabc9061. 10.1126/sciadv.abc9061.

[156] Dhondt B, Pinheiro C, Geeurickx E, Tulkens J, Vergauwen G, Van Der Pol E, Nieuwland R, Decock A, Miinalainen I, Rappu P, Schroth G, Kuersten S, Vandesompele J, Mestdagh P, Lumen N, De Wever O, Hendrix A (2023) Benchmarking blood collection tubes and processing intervals for extracellular vesicle performance metrics. J Extracell Vesicles 12, e12315. 10.1002/jev2.12315.

[157] Handtke S, Thiele T (2020) Large and small platelets-(When) do they differ? J Thromb Haemost 18, 1256–1267. 10.1111/jth.14788.

[158] Bracht JWP, Los M, van Eijndhoven MAJ, Bettin B, van der Pol E, Pegtel DM, Nieuwland R (2023) Platelet removal from human blood plasma improves detection of extracellular vesicle-associated miRNA. J Extracell Vesicles 12, e12302. 10.1002/jev2.12302.

[159] Patel GK, Khan MA, Zubair H, Srivastava SK, Khushman M, Singh S, Singh AP (2019) Comparative analysis of exosome isolation methods using culture supernatant for optimum yield, purity and downstream applications. Sci Rep 9, 5335. 10.1038/s41598-019-41800-2.

[160] Hendrix A, Lippens L, Pinheiro C, Théry C, Martin-Jaular L, Lötvall J, Lässer C, Hill A, Witwer K (2023) Extracellular vesicle analysis. Nat Rev Dis Primers 3, 56. 10.1038/s43586-023-00240-z.

[161] Gelibter S, Marostica G, Mandelli A, Siciliani S, Podini P, Finardi A, Furlan R (2022) The impact of storage on extracellular vesicles: A systematic study. J Extracell Vesicles 11, e12162. 10.1002/jev2.12162.

[162] Gomes DE, Witwer KW (2022) L1CAM-associated extracellular vesicles: A systematic review of nomenclature, sources, separation, and characterization. J Extracell Biol 1. 10.1002/jex2.35.

[163] Ter-Ovanesyan D, Whiteman S, Gilboa T, Kowal EJK, Trieu W, Lyer S, Budnik B, Babila CM, Heimberg G, Burgess MW, Keshishian H, Carr SA, Regev A, Church GM, Walt DR (2024) Identification of markers for the isolation of neuron-specific extracellular vesicles. bioRxiv. 10.1101/2024.04.03.587267.

[164] Hosoki S, Hansra GK, Jayasena T, Poljak A, Mather KA, Catts VS, Rust R, Sagare A, Kovacic JC, Brodtmann A, Wallin A, Zlokovic BV, Ihara M, Sachdev PS (2023) Molecular biomarkers for vascular cognitive impairment and dementia. Nat Rev Neurol 19, 737–753. 10.1038/s41582-023-00884-1.

[165] Foley KE, Wilcock DM (2024) Soluble Biomarkers of Cerebrovascular Pathologies. Stroke 55, 801–811. 10.1161/STROKEAHA.123.044172.

[166] Buchholz S, Zempel H (2024) The six brain-specific TAU isoforms and their role in Alzheimer’s disease and related neurodegenerative dementia syndromes. Alzheimers Dement. 10.1002/alz.13784.

[167] Kumar A, Sidhu J, Goyal A, Tsao JW (2024) Alzheimer Disease In StatPearls, Treasure Island (FL).

[168] Barthelemy NR, Bateman RJ, Hirtz C, Marin P, Becher F, Sato C, Gabelle A, Lehmann S (2020) Cerebrospinal fluid phospho-tau T217 outperforms T181 as a biomarker for the differential diagnosis of Alzheimer’s disease and PET amyloid-positive patient identification. Alzheimers Res Ther 12, 26. 10.1186/s13195-020-00596-4.

[169] Ashton NJ, Brum WS, Di Molfetta G, Benedet AL, Arslan B, Jonaitis E, Langhough RE, Cody K, Wilson R, Carlsson CM, Vanmechelen E, Montoliu-Gaya L, Lantero-Rodriguez J, Rahmouni N, Tissot C, Stevenson J, Servaes S, Therriault J, Pascoal T, Lleo A, Alcolea D, Fortea J, Rosa-Neto P, Johnson S, Jeromin A, Blennow K, Zetterberg H (2024) Diagnostic Accuracy of a Plasma Phosphorylated Tau 217 Immunoassay for Alzheimer Disease Pathology. JAMA Neurol. 10.1001/jamaneurol.2023.5319.

[170] Gonzalez-Ortiz F, Ferreira PCL, Gonzalez-Escalante A, Montoliu-Gaya L, Ortiz-Romero P, Kac PR, Turton M, Kvartsberg H, Ashton NJ, Zetterberg H, Harrison P, Bellaver B, Povala G, Villemagne VL, Pascoal TA, Ganguli M, Cohen AD, Minguillon C, Contador J, Suarez-Calvet M, Karikari TK, Blennow K (2023) A novel ultrasensitive assay for plasma p-tau217: Performance in individuals with subjective cognitive decline and early Alzheimer’s disease. Alzheimers Dement. 10.1002/alz.13525.

[171] Taha HB (2024) Early detection of subjective cognitive decline and Alzheimer’s disease: Analytical validation of a newly developed pT217-tau assay. Alzheimer’s and Dementia. 10.1002/alz.13707.

[172] Janelidze S, Bali D, Ashton NJ, Barthelemy NR, Vanbrabant J, Stoops E, Vanmechelen E, He Y, Dolado AO, Triana-Baltzer G, Pontecorvo MJ, Zetterberg H, Kolb H, Vandijck M, Blennow K, Bateman RJ, Hansson O (2023) Head-to-head comparison of 10 plasma phospho-tau assays in prodromal Alzheimer’s disease. Brain 146, 1592–1601. 10.1093/brain/awac333.

[173] Feizpour A, Dore V, Doecke JD, Saad ZS, Triana-Baltzer G, Slemmon R, Maruff P, Krishnadas N, Bourgeat P, Huang K, Fowler C, Rainey-Smith SR, Bush AI, Ward L, Robertson J, Martins RN, Masters CL, Villemagne VL, Fripp J, Kolb HC, Rowe CC (2023) Two-Year Prognostic Utility of Plasma p217+tau across the Alzheimer’s Continuum. J Prev Alzheimers Dis 10, 828–836. 10.14283/jpad.2023.83.

[174] Palmqvist S, Janelidze S, Quiroz YT, Zetterberg H, Lopera F, Stomrud E, Su Y, Chen Y, Serrano GE, Leuzy A, Mattsson-Carlgren N, Strandberg O, Smith R, Villegas A, Sepulveda-Falla D, Chai X, Proctor NK, Beach TG, Blennow K, Dage JL, Reiman EM, Hansson O (2020) Discriminative Accuracy of Plasma Phospho-tau217 for Alzheimer Disease vs Other Neurodegenerative Disorders. JAMA 324, 772–781. 10.1001/jama.2020.12134.

[175] Gobom J, Benedet AL, Mattsson-Carlgren N, Montoliu-Gaya L, Schultz N, Ashton NJ, Janelidze S, Servaes S, Sauer M, Pascoal TA, Karikari TK, Lantero-Rodriguez J, Brinkmalm G, Zetterberg H, Hansson O, Rosa-Neto P, Blennow K (2022) Antibody-free measurement of cerebrospinal fluid tau phosphorylation across the Alzheimer’s disease continuum. Mol Neurodegener 17, 81. 10.1186/s13024-022-00586-0.

[176] Zhang D, Zhang W, Ming C, Gao X, Yuan H, Lin X, Mao X, Wang C, Guo X, Du Y, Shao L, Yang R, Lin Z, Wu X, Huang TY, Wang Z, Zhang YW, Xu H, Zhao Y (2024) P-tau217 correlates with neurodegeneration in Alzheimer’s disease, and targeting p-tau217 with immunotherapy ameliorates murine tauopathy. Neuron. 10.1016/j.neuron.2024.02.017.

[177] Bernal-Conde LD, Ramos-Acevedo R, Reyes-Hernandez MA, Balbuena-Olvera AJ, Morales-Moreno ID, Arguero-Sanchez R, Schule B, Guerra-Crespo M (2019) Alpha-Synuclein Physiology and Pathology: A Perspective on Cellular Structures and Organelles. Front Neurosci 13, 1399. 10.3389/fnins.2019.01399.

[178] Taha HB, Hornung S, Dutta S, Fenwick L, Lahgui O, Howe K, Elabed N, Del Rosario I, Wong DY, Duarte Folle A, Markovic D, Palma JA, Kang UJ, Alcalay RN, Sklerov M, Kaufmann H, Fogel BL, Bronstein JM, Ritz B, Bitan G (2023) Toward a biomarker panel measured in CNS-originating extracellular vesicles for improved differential diagnosis of Parkinson’s disease and multiple system atrophy. Transl Neurodegener 12, 14. 10.1186/s40035-023-00346-0.

[179] Espay AJ, Lees AJ (2024) Are we entering the ‘Tau-lemaic’ era of Parkinson’s disease? Brain 147, 330–332. 10.1093/brain/awae002.

[180] Zhang X, Gao F, Wang D, Li C, Fu Y, He W, Zhang J (2018) Tau Pathology in Parkinson’s Disease. Front Neurol 9, 809. 10.3389/fneur.2018.00809.

[181] Chu Y, Hirst WD, Federoff HJ, Harms AS, Stoessl AJ, Kordower JH (2024) Nigrostriatal tau pathology in parkinsonism and Parkinson’s disease. Brain 147, 444–457. 10.1093/brain/awad388.

[182] Nagaishi M, Yokoo H, Nakazato Y (2011) Tau-positive glial cytoplasmic granules in multiple system atrophy. Neuropathology 31, 299–305. 10.1111/j.1440-1789.2010.01159.x.

[183] Campese N, Fanciulli A, Stefanova N, Haybaeck J, Kiechl S, Wenning GK (2021) Neuropathology of multiple system atrophy: Kurt Jellinger;s legacy. J Neural Transm (Vienna) 128, 1481–1494. 10.1007/s00702-021-02383-3.

[184] Bujan B, Hofer MJ, Oertel WH, Pagenstecher A, Burk K (2013) Multiple system atrophy of the cerebellar type (MSA-C) with concomitant beta-amyloid and tau pathology. Clin Neuropathol 32, 286–290. 10.5414/NP300572.

[185] Tolosa E, Garrido A, Scholz SW, Poewe W (2021) Challenges in the diagnosis of Parkinson’s disease. Lancet Neurol 20, 385–397. 10.1016/S1474-4422(21)00030-2.

[186] Poewe W, Stankovic I, Halliday G, Meissner WG, Wenning GK, Pellecchia MT, Seppi K, Palma JA, Kaufmann H (2022) Multiple system atrophy. Nat Rev Dis Primers 8, 56. 10.1038/s41572-022-00382-6.

[187] Postuma RB, Berg D, Stern M, Poewe W, Olanow CW, Oertel W, Obeso J, Marek K, Litvan I, Lang AE, Halliday G, Goetz CG, Gasser T, Dubois B, Chan P, Bloem BR, Adler CH, Deuschl G (2015) MDS clinical diagnostic criteria for Parkinson’s disease. Mov Disord 30, 1591–1601. 10.1002/mds.26424.

[188] Gilman S, Wenning GK, Low PA, Brooks DJ, Mathias CJ, Trojanowski JQ, Wood NW, Colosimo C, Durr A, Fowler CJ, Kaufmann H, Klockgether T, Lees A, Poewe W, Quinn N, Revesz T, Robertson D, Sandroni P, Seppi K, Vidailhet M (2008) Second consensus statement on the diagnosis of multiple system atrophy. Neurology 71, 670–676. 10.1212/01.wnl.0000324625.00404.15.

[189] Miglis MG, Adler CH, Antelmi E, Arnaldi D, Baldelli L, Boeve BF, Cesari M, Dall’Antonia I, Diederich NJ, Doppler K, Dusek P, Ferri R, Gagnon JF, Gan-Or Z, Hermann W, Hogl B, Hu MT, Iranzo A, Janzen A, Kuzkina A, Lee JY, Leenders KL, Lewis SJG, Liguori C, Liu J, Lo C, Ehgoetz Martens KA, Nepozitek J, Plazzi G, Provini F, Puligheddu M, Rolinski M, Rusz J, Stefani A, Summers RLS, Yoo D, Zitser J, Oertel WH (2021) Biomarkers of conversion to alpha-synucleinopathy in isolated rapid-eye-movement sleep behaviour disorder. Lancet Neurol 20, 671–684. 10.1016/S1474-4422(21)00176-9.

[190] Li T, Le W (2020) Biomarkers for Parkinson’s Disease: How Good Are They? Neurosci Bull 36, 183–194. 10.1007/s12264-019-00433-1.

[191] Taha HB, Ati SA (2023) Evaluation of α-synuclein in CNS-originating extracellular vesicles for Parkinsonian disorders: A systematic review and meta-analysis. CNS Neurosci Ther, 1–15. 10.1111/cns.14341.

[192] Taha HB (2024) Biomarker bust: meta-analyses reveal unreliability of neuronal extracellular vesicles for diagnosing parkinsonian disorders. Neural Regen Res. 10.4103/NRR.NRR-D-23-02102.

[193] Concha-Marambio L, Pritzkow S, Shahnawaz M, Farris CM, Soto C (2023) Seed amplification assay for the detection of pathologic alpha-synuclein aggregates in cerebrospinal fluid. Nat Protoc 18, 1179–1196. 10.1038/s41596-022-00787-3.

[194] Fernandes Gomes B, Farris CM, Ma Y, Concha-Marambio L, Lebovitz R, Nellgard B, Dalla K, Constantinescu J, Constantinescu R, Gobom J, Andreasson U, Zetterberg H, Blennow K (2023) alpha-Synuclein seed amplification assay as a diagnostic tool for parkinsonian disorders. Parkinsonism Relat Disord 117, 105807. 10.1016/j.parkreldis.2023.105807.

[195] Bellomo G, De Luca CMG, Paoletti FP, Gaetani L, Moda F, Parnetti L (2022) alpha-Synuclein Seed Amplification Assays for Diagnosing Synucleinopathies: The Way Forward. Neurology 99, 195–205. 10.1212/WNL.0000000000200878.

[196] Grossauer A, Hemicker G, Krismer F, Peball M, Djamshidian A, Poewe W, Seppi K, Heim B (2023) alpha-Synuclein Seed Amplification Assays in the Diagnosis of Synucleinopathies Using Cerebrospinal Fluid-A Systematic Review and Meta-Analysis. Mov Disord Clin Pract 10, 737–747. 10.1002/mdc3.13710.

[197] Yoo D, Bang JI, Ahn C, Nyaga VN, Kim YE, Kang MJ, Ahn TB (2022) Diagnostic value of alpha-synuclein seeding amplification assays in alpha-synucleinopathies: A systematic review and meta-analysis. Parkinsonism Relat Disord 104, 99–109. 10.1016/j.parkreldis.2022.10.007.

[198] Ferreira NDC, Caughey B (2020) Proteopathic Seed Amplification Assays for Neurodegenerative Disorders. Clin Lab Med 40, 257–270. 10.1016/j.cll.2020.04.002.

[199] Concha-Marambio L, Farris CM, Holguin B, Ma Y, Seibyl J, Russo MJ, Kang UJ, Hutten SJ, Merchant K, Shahnawaz M, Soto C (2021) Seed Amplification Assay to Diagnose Early Parkinson’s and Predict Dopaminergic Deficit Progression. Mov Disord 36, 2444–2446. 10.1002/mds.28715.

[200] Russo MJ, Orru CD, Concha-Marambio L, Giaisi S, Groveman BR, Farris CM, Holguin B, Hughson AG, LaFontant DE, Caspell-Garcia C, Coffey CS, Mollon J, Hutten SJ, Merchant K, Heym RG, Soto C, Caughey B, Kang UJ (2021) High diagnostic performance of independent alpha-synuclein seed amplification assays for detection of early Parkinson’s disease. Acta Neuropathol Commun 9, 179. 10.1186/s40478-021-01282-8.

[201] Chahine LM, Beach TG, Adler CH, Hepker M, Kanthasamy A, Appel S, Pritzkow S, Pinho M, Mosovsky S, Serrano GE, Coffey C, Brumm MC, Oliveira LMA, Eberling J, Mollenhauer B, Systemic Synuclein Sampling S (2023) Central and peripheral alpha-synuclein in Parkinson disease detected by seed amplification assay. Ann Clin Transl Neurol 10, 696–705. 10.1002/acn3.51753.

[202] Middleton JS, Hovren HL, Kha N, Medina MJ, MacLeod KR, Concha-Marambio L, Jensen KJ (2023) Seed amplification assay results illustrate discrepancy in Parkinson’s disease clinical diagnostic accuracy and error rates. J Neurol 270, 5813–5818. 10.1007/s00415-023-11810-2.

[203] Kluge A, Bunk J, Schaeffer E, Drobny A, Xiang W, Knacke H, Bub S, Lückstädt W, Arnold P, Lucius R, Berg D, Zunke F (2022) Detection of neuron-derived pathological α-synuclein in blood. Brain 145, 3058–3071. 10.1093/brain/awac115.

[204] Brauer S, Rossi M, Sajapin J, Henle T, Gasser T, Parchi P, Brockmann K, Falkenburger BH (2023) Kinetic parameters of alpha-synuclein seed amplification assay correlate with cognitive impairment in patients with Lewy body disorders. Acta Neuropathol Commun 11, 162. 10.1186/s40478-023-01653-3.

[205] Kluge A, Borsche M, Streubel-Gallasch L, Gul T, Schaake S, Balck A, Prasuhn J, Campbell P, Morris HR, Schapira AH, Lohmann K, Bruggemann N, Rakovic A, Seibler P, Basak AN, Berg D, Klein C (2024) alpha-Synuclein Pathology in PRKN-Linked Parkinson’s Disease: New Insights from a Blood-Based Seed Amplification Assay. Ann Neurol. 10.1002/ana.26917.

[206] Matthews I, Birnbaum A, Gromova A, Huang AW, Liu K, Liu EA, Coutinho K, McGraw M, Patterson DC, Banks MT, Nobles AC, Nguyen N, Merrihew GE, Wang L, Baeuerle E, Fernandez E, Musi N, MacCoss MJ, Miranda HC, La Spada AR, Cortes CJ (2023) Skeletal muscle TFEB signaling promotes central nervous system function and reduces neuroinflammation during aging and neurodegenerative disease. Cell Rep 42, 113436. 10.1016/j.celrep.2023.113436.

[207] Gupta R, Khan R, Cortes CJ (2021) Forgot to Exercise? Exercise Derived Circulating Myokines in Alzheimer’s Disease: A Perspective. Front Neurol 12, 649452. 10.3389/fneur.2021.649452.

[208] Nederveen JP, Warnier G, Di Carlo A, Nilsson MI, Tarnopolsky MA (2020) Extracellular Vesicles and Exosomes: Insights From Exercise Science. Front Physiol 11, 604274. 10.3389/fphys.2020.604274.

[209] Pierdona TM, Martin A, Obi PO, Seif S, Bydak B, Labouta HI, Eadie AL, Brunt KR, McGavock JM, Senechal M, Saleem A (2022) Extracellular Vesicles as Predictors of Individual Response to Exercise Training in Youth Living with Obesity. Front Biosci (Landmark Ed) 27, 143. 10.31083/j.fbl2705143.

[210] Lim AC, Barnes LL, Weissberger GH, Lamar M, Nguyen AL, Fenton L, Herrera J, Han SD (2023) Quantification of race/ethnicity representation in Alzheimer’s disease neuroimaging research in the USA: a systematic review. Commun Med (Lond) 3, 101. 10.1038/s43856-023-00333-6.

[211] Deters KD, Mormino EC, Yu L, Lutz MW, Bennett DA, Barnes LL (2021) TOMM40-APOE haplotypes are associated with cognitive decline in non-demented Blacks. Alzheimers Dement 17, 1287–1296. 10.1002/alz.12295.

[212] Deters KD, Napolioni V, Sperling RA, Greicius MD, Mayeux R, Hohman T, Mormino EC (2021) Amyloid PET Imaging in Self-Identified Non-Hispanic Black Participants of the Anti-Amyloid in Asymptomatic Alzheimer’s Disease (A4) Study. Neurology 96, e1491–e1500. 10.1212/WNL.0000000000011599.

[213] Arce Renteria M, Mobley TM, Evangelista ND, Medina LD, Deters KD, Fox-Fuller JT, Minto LR, Avila-Rieger J, Bettcher BM (2023) Representativeness of samples enrolled in Alzheimer’s disease research centers. Alzheimers Dement (Amst) 15, e12450. 10.1002/dad2.12450.

[214] Barnes LL (2019) Biomarkers for Alzheimer Dementia in Diverse Racial and Ethnic Minorities-A Public Health Priority. JAMA Neurol 76, 251–253. 10.1001/jamaneurol.2018.3444.

[215] Manly JJ, Gilmore-Bykovskyi A, Deters KD (2021) Inclusion of Underrepresented Groups in Preclinical Alzheimer Disease Trials-Opportunities Abound. JAMA Netw Open 4, e2114606. 10.1001/jamanetworkopen.2021.14606.

[216] Turney IC, Lao PJ, Renteria MA, Igwe KC, Berroa J, Rivera A, Benavides A, Morales CD, Rizvi B, Schupf N, Mayeux R, Manly JJ, Brickman AM (2023) Brain Aging Among Racially and Ethnically Diverse Middle-Aged and Older Adults. JAMA Neurol 80, 73–81. 10.1001/jamaneurol.2022.3919.

[217] Sandau US, Magana SM, Costa J, Nolan JP, Ikezu T, Vella LJ, Jackson HK, Moreira LR, Palacio PL, Hill AF, Quinn JF, Van Keuren-Jensen KR, McFarland TJ, Palade J, Sribnick EA, Su H, Vekrellis K, Coyle B, Yang Y, Falcon-Perez JM, Nieuwland R, Saugstad JA, International Society for Extracellular Vesicles Cerebrospinal Fluid Task F (2024) Recommendations for reproducibility of cerebrospinal fluid extracellular vesicle studies. J Extracell Vesicles 13, e12397. 10.1002/jev2.12397.

[218] Lucien F, Gustafson D, Lenassi M, Li B, Teske JJ, Boilard E, von Hohenberg KC, Falcon-Perez JM, Gualerzi A, Reale A, Jones JC, Lasser C, Lawson C, Nazarenko I, O’Driscoll L, Pink R, Siljander PR, Soekmadji C, Hendrix A, Welsh JA, Witwer KW, Nieuwland R (2023) MIBlood-EV: Minimal information to enhance the quality and reproducibility of blood extracellular vesicle research. J Extracell Vesicles 12, e12385. 10.1002/jev2.12385.

[219] Clayton A, Boilard E, Buzas EI, Cheng L, Falcon-Perez JM, Gardiner C, Gustafson D, Gualerzi A, Hendrix A, Hoffman A, Jones J, Lasser C, Lawson C, Lenassi M, Nazarenko I, O’Driscoll L, Pink R, Siljander PR, Soekmadji C, Wauben M, Welsh JA, Witwer K, Zheng L, Nieuwland R (2019) Considerations towards a roadmap for collection, handling and storage of blood extracellular vesicles. J Extracell Vesicles 8, 1647027. 10.1080/20013078.2019.1647027.

[220] van Royen ME, Soekmadji C, Grange C, Webber JP, Tertel T, Droste M, Buescher A, Giebel B, Jenster GW, Llorente A, Blijdorp CJ, Burger D, Erdbrugger U, Martens-Uzunova ES (2023) The quick reference card “Storage of urinary EVs” – A practical guideline tool for research and clinical laboratories. J Extracell Vesicles 12, e12286. 10.1002/jev2.12286.

[221] Blijdorp CJ, Burger D, Llorente A, Martens-Uzunova ES, Erdbrugger U (2022) Extracellular Vesicles as Novel Players in Kidney Disease. J Am Soc Nephrol 33, 467–471. 10.1681/ASN.2021091232.

[222] Erdbrugger U, Blijdorp CJ, Bijnsdorp IV, Borras FE, Burger D, Bussolati B, Byrd JB, Clayton A, Dear JW, Falcon-Perez JM, Grange C, Hill AF, Holthofer H, Hoorn EJ, Jenster G, Jimenez CR, Junker K, Klein J, Knepper MA, Koritzinsky EH, Luther JM, Lenassi M, Leivo J, Mertens I, Musante L, Oeyen E, Puhka M, van Royen ME, Sanchez C, Soekmadji C, Thongboonkerd V, van Steijn V, Verhaegh G, Webber JP, Witwer K, Yuen PST, Zheng L, Llorente A, Martens-Uzunova ES (2021) Urinary extracellular vesicles: A position paper by the Urine Task Force of the International Society for Extracellular Vesicles. J Extracell Vesicles 10, e12093. 10.1002/jev2.12093.

[223] Lötvall J, Hill AF, Hochberg F, Buzas EI, Di Vizio D, Gardiner C, Gho YS, Kurochkin IV, Mathivanan S, Quesenberry P, Sahoo S, Tahara H, Wauben MH, Witwer KW, Thery C (2014) Minimal experimental requirements for definition of extracellular vesicles and their functions: a position statement from the International Society for Extracellular Vesicles. J Extracell Vesicles 3, 26913. 10.3402/jev.v3.26913.

[224] Thery C, Witwer KW, Aikawa E, Alcaraz MJ, Anderson JD, Andriantsitohaina R, Antoniou A, Arab T, Archer F, Atkin-Smith GK, Ayre DC, Bach JM, Bachurski D, Baharvand H, Balaj L, Baldacchino S, Bauer NN, Baxter AA, Bebawy M, Beckham C, Bedina Zavec A, Benmoussa A, Berardi AC, Bergese P, Bielska E, Blenkiron C, Bobis-Wozowicz S, Boilard E, Boireau W, Bongiovanni A, Borras FE, Bosch S, Boulanger CM, Breakefield X, Breglio AM, Brennan MA, Brigstock DR, Brisson A, Broekman ML, Bromberg JF, Bryl-Gorecka P, Buch S, Buck AH, Burger D, Busatto S, Buschmann D, Bussolati B, Buzas EI, Byrd JB, Camussi G, Carter DR, Caruso S, Chamley LW, Chang YT, Chen C, Chen S, Cheng L, Chin AR, Clayton A, Clerici SP, Cocks A, Cocucci E, Coffey RJ, Cordeiro-da-Silva A, Couch Y, Coumans FA, Coyle B, Crescitelli R, Criado MF, D’Souza-Schorey C, Das S, Datta Chaudhuri A, de Candia P, De Santana EF, De Wever O, Del Portillo HA, Demaret T, Deville S, Devitt A, Dhondt B, Di Vizio D, Dieterich LC, Dolo V, Dominguez Rubio AP, Dominici M, Dourado MR, Driedonks TA, Duarte FV, Duncan HM, Eichenberger RM, Ekstrom K, El Andaloussi S, Elie-Caille C, Erdbrugger U, Falcon-Perez JM, Fatima F, Fish JE, Flores-Bellver M, Forsonits A, Frelet-Barrand A, Fricke F, Fuhrmann G, Gabrielsson S, Gamez-Valero A, Gardiner C, Gartner K, Gaudin R, Gho YS, Giebel B, Gilbert C, Gimona M, Giusti I, Goberdhan DC, Gorgens A, Gorski SM, Greening DW, Gross JC, Gualerzi A, Gupta GN, Gustafson D, Handberg A, Haraszti RA, Harrison P, Hegyesi H, Hendrix A, Hill AF, Hochberg FH, Hoffmann KF, Holder B, Holthofer H, Hosseinkhani B, Hu G, Huang Y, Huber V, Hunt S, Ibrahim AG, Ikezu T, Inal JM, Isin M, Ivanova A, Jackson HK, Jacobsen S, Jay SM, Jayachandran M, Jenster G, Jiang L, Johnson SM, Jones JC, Jong A, Jovanovic-Talisman T, Jung S, Kalluri R, Kano SI, Kaur S, Kawamura Y, Keller ET, Khamari D, Khomyakova E, Khvorova A, Kierulf P, Kim KP, Kislinger T, Klingeborn M, Klinke DJ, 2nd, Kornek M, Kosanovic MM, Kovacs AF, Kramer-Albers EM, Krasemann S, Krause M, Kurochkin IV, Kusuma GD, Kuypers S, Laitinen S, Langevin SM, Languino LR, Lannigan J, Lasser C, Laurent LC, Lavieu G, Lazaro-Ibanez E, Le Lay S, Lee MS, Lee YXF, Lemos DS, Lenassi M, Leszczynska A, Li IT, Liao K, Libregts SF, Ligeti E, Lim R, Lim SK, Line A, Linnemannstons K, Llorente A, Lombard CA, Lorenowicz MJ, Lorincz AM, Lotvall J, Lovett J, Lowry MC, Loyer X, Lu Q, Lukomska B, Lunavat TR, Maas SL, Malhi H, Marcilla A, Mariani J, Mariscal J, Martens-Uzunova ES, Martin-Jaular L, Martinez MC, Martins VR, Mathieu M, Mathivanan S, Maugeri M, McGinnis LK, McVey MJ, Meckes DG, Jr., Meehan KL, Mertens I, Minciacchi VR, Moller A, Moller Jorgensen M, Morales-Kastresana A, Morhayim J, Mullier F, Muraca M, Musante L, Mussack V, Muth DC, Myburgh KH, Najrana T, Nawaz M, Nazarenko I, Nejsum P, Neri C, Neri T, Nieuwland R, Nimrichter L, Nolan JP, Nolte-’t Hoen EN, Noren Hooten N, O’Driscoll L, O’Grady T, O’Loghlen A, Ochiya T, Olivier M, Ortiz A, Ortiz LA, Osteikoetxea X, Ostergaard O, Ostrowski M, Park J, Pegtel DM, Peinado H, Perut F, Pfaffl MW, Phinney DG, Pieters BC, Pink RC, Pisetsky DS, Pogge von Strandmann E, Polakovicova I, Poon IK, Powell BH, Prada I, Pulliam L, Quesenberry P, Radeghieri A, Raffai RL, Raimondo S, Rak J, Ramirez MI, Raposo G, Rayyan MS, Regev-Rudzki N, Ricklefs FL, Robbins PD, Roberts DD, Rodrigues SC, Rohde E, Rome S, Rouschop KM, Rughetti A, Russell AE, Saa P, Sahoo S, Salas-Huenuleo E, Sanchez C, Saugstad JA, Saul MJ, Schiffelers RM, Schneider R, Schoyen TH, Scott A, Shahaj E, Sharma S, Shatnyeva O, Shekari F, Shelke GV, Shetty AK, Shiba K, Siljander PR, Silva AM, Skowronek A, Snyder OL, 2nd, Soares RP, Sodar BW, Soekmadji C, Sotillo J, Stahl PD, Stoorvogel W, Stott SL, Strasser EF, Swift S, Tahara H, Tewari M, Timms K, Tiwari S, Tixeira R, Tkach M, Toh WS, Tomasini R, Torrecilhas AC, Tosar JP, Toxavidis V, Urbanelli L, Vader P, van Balkom BW, van der Grein SG, Van Deun J, van Herwijnen MJ, Van Keuren-Jensen K, van Niel G, van Royen ME, van Wijnen AJ, Vasconcelos MH, Vechetti IJ, Jr., Veit TD, Vella LJ, Velot E, Verweij FJ, Vestad B, Vinas JL, Visnovitz T, Vukman KV, Wahlgren J, Watson DC, Wauben MH, Weaver A, Webber JP, Weber V, Wehman AM, Weiss DJ, Welsh JA, Wendt S, Wheelock AM, Wiener Z, Witte L, Wolfram J, Xagorari A, Xander P, Xu J, Yan X, Yanez-Mo M, Yin H, Yuana Y, Zappulli V, Zarubova J, Zekas V, Zhang JY, Zhao Z, Zheng L, Zheutlin AR, Zickler AM, Zimmermann P, Zivkovic AM, Zocco D, Zuba-Surma EK (2018) Minimal information for studies of extracellular vesicles 2018 (MISEV2018): a position statement of the International Society for Extracellular Vesicles and update of the MISEV2014 guidelines. J Extracell Vesicles 7, 1535750. 10.1080/20013078.2018.1535750.

[225] Elahi FM, Miller BL (2017) A clinicopathological approach to the diagnosis of dementia. Nat Rev Neurol 13, 457–476. 10.1038/nrneurol.2017.96.

[226] Roberts RO, Aakre JA, Kremers WK, Vassilaki M, Knopman DS, Mielke MM, Alhurani R, Geda YE, Machulda MM, Coloma P, Schauble B, Lowe VJ, Jack CR, Jr., Petersen RC (2018) Prevalence and Outcomes of Amyloid Positivity Among Persons Without Dementia in a Longitudinal, Population-Based Setting. JAMA Neurol 75, 970–979. 10.1001/jamaneurol.2018.0629.

[227] Ossenkoppele R, Rabinovici GD, Smith R, Cho H, Scholl M, Strandberg O, Palmqvist S, Mattsson N, Janelidze S, Santillo A, Ohlsson T, Jogi J, Tsai R, La Joie R, Kramer J, Boxer AL, Gorno-Tempini ML, Miller BL, Choi JY, Ryu YH, Lyoo CH, Hansson O (2018) Discriminative Accuracy of [18F]flortaucipir Positron Emission Tomography for Alzheimer Disease vs Other Neurodegenerative Disorders. JAMA 320, 1151–1162. 10.1001/jama.2018.12917.

[228] Eckerstrom C, Svensson J, Kettunen P, Jonsson M, Eckerstrom M (2021) Evaluation of the ATN model in a longitudinal memory clinic sample with different underlying disorders. Alzheimers Dement (Amst) 13, e12031. 10.1002/dad2.12031.

[229] Jansen WJ, Ossenkoppele R, Knol DL, Tijms BM, Scheltens P, Verhey FR, Visser PJ, Amyloid Biomarker Study G, Aalten P, Aarsland D, Alcolea D, Alexander M, Almdahl IS, Arnold SE, Baldeiras I, Barthel H, van Berckel BN, Bibeau K, Blennow K, Brooks DJ, van Buchem MA, Camus V, Cavedo E, Chen K, Chetelat G, Cohen AD, Drzezga A, Engelborghs S, Fagan AM, Fladby T, Fleisher AS, van der Flier WM, Ford L, Forster S, Fortea J, Foskett N, Frederiksen KS, Freund-Levi Y, Frisoni GB, Froelich L, Gabryelewicz T, Gill KD, Gkatzima O, Gomez-Tortosa E, Gordon MF, Grimmer T, Hampel H, Hausner L, Hellwig S, Herukka SK, Hildebrandt H, Ishihara L, Ivanoiu A, Jagust WJ, Johannsen P, Kandimalla R, Kapaki E, Klimkowicz-Mrowiec A, Klunk WE, Kohler S, Koglin N, Kornhuber J, Kramberger MG, Van Laere K, Landau SM, Lee DY, de Leon M, Lisetti V, Lleo A, Madsen K, Maier W, Marcusson J, Mattsson N, de Mendonca A, Meulenbroek O, Meyer PT, Mintun MA, Mok V, Molinuevo JL, Mollergard HM, Morris JC, Mroczko B, Van der Mussele S, Na DL, Newberg A, Nordberg A, Nordlund A, Novak GP, Paraskevas GP, Parnetti L, Perera G, Peters O, Popp J, Prabhakar S, Rabinovici GD, Ramakers IH, Rami L, Resende de Oliveira C, Rinne JO, Rodrigue KM, Rodriguez-Rodriguez E, Roe CM, Rot U, Rowe CC, Ruther E, Sabri O, Sanchez-Juan P, Santana I, Sarazin M, Schroder J, Schutte C, Seo SW, Soetewey F, Soininen H, Spiru L, Struyfs H, Teunissen CE, Tsolaki M, Vandenberghe R, Verbeek MM, Villemagne VL, Vos SJ, van Waalwijk van Doorn LJ, Waldemar G, Wallin A, Wallin AK, Wiltfang J, Wolk DA, Zboch M, Zetterberg H (2015) Prevalence of cerebral amyloid pathology in persons without dementia: a meta-analysis. JAMA 313, 1924–1938. 10.1001/jama.2015.4668.

[230] Jack CR, Jr., Bennett DA, Blennow K, Carrillo MC, Feldman HH, Frisoni GB, Hampel H, Jagust WJ, Johnson KA, Knopman DS, Petersen RC, Scheltens P, Sperling RA, Dubois B (2016) A/T/N: An unbiased descriptive classification scheme for Alzheimer disease biomarkers. Neurology 87, 539–547. 10.1212/WNL.0000000000002923.

[231] Korczyn AD, Grinberg LT (2024) Is Alzheimer disease a disease? Nat Rev Neurol 20, 245–251. 10.1038/s41582-024-00940-4.

[232] Shelton JF, Geraghty EM, Tancredi DJ, Delwiche LD, Schmidt RJ, Ritz B, Hansen RL, Hertz-Picciotto I (2014) Neurodevelopmental disorders and prenatal residential proximity to agricultural pesticides: the CHARGE study. Environ Health Perspect 122, 1103–1109. 10.1289/ehp.1307044.

[233] Haghani A, Cacciottolo M, Doty KR, D’Agostino C, Thorwald M, Safi N, Levine ME, Sioutas C, Town TC, Forman HJ, Zhang H, Morgan TE, Finch CE (2020) Mouse brain transcriptome responses to inhaled nanoparticulate matter differed by sex and APOE in Nrf2-Nfkb interactions. Elife 9. 10.7554/eLife.54822.

[234] Finch CE (2023) Air pollution, dementia, and lifespan in the socio-economic gradient of aging: perspective on human aging for planning future experimental studies. Front Aging 4, 1273303. 10.3389/fragi.2023.1273303.

[235] Shkirkova K, Lamorie-Foote K, Zhang N, Li A, Diaz A, Liu Q, Thorwald MA, Godoy-Lugo JA, Ge B, D’Agostino C, Zhang Z, Mack WJ, Sioutas C, Finch CE, Mack WJ, Zhang H (2022) Neurotoxicity of Diesel Exhaust Particles. J Alzheimers Dis 89, 1263–1278. 10.3233/JAD-220493.

[236] Dugger BN, Dickson DW (2017) Pathology of Neurodegenerative Diseases. Cold Spring Harb Perspect Biol 9. 10.1101/cshperspect.a028035.

[237] Ward A, Tardiff S, Dye C, Arrighi HM (2013) Rate of conversion from prodromal Alzheimer’s disease to Alzheimer’s dementia: a systematic review of the literature. Dement Geriatr Cogn Dis Extra 3, 320–332. 10.1159/000354370.

[238] Shigemizu D, Akiyama S, Higaki S, Sugimoto T, Sakurai T, Boroevich KA, Sharma A, Tsunoda T, Ochiya T, Niida S, Ozaki K (2020) Prognosis prediction model for conversion from mild cognitive impairment to Alzheimer’s disease created by integrative analysis of multi-omics data. Alzheimers Res Ther 12, 145. 10.1186/s13195-020-00716-0.

[239] Chen Y, Qian X, Zhang Y, Su W, Huang Y, Wang X, Chen X, Zhao E, Han L, Ma Y (2022) Prediction Models for Conversion From Mild Cognitive Impairment to Alzheimer’s Disease: A Systematic Review and Meta-Analysis. Front Aging Neurosci 14, 840386. 10.3389/fnagi.2022.840386.

[240] Fierz W (2018) Likelihood ratios of quantitative laboratory results in medical diagnosis: The application of Bezier curves in ROC analysis. PLoS One 13, e0192420. 10.1371/journal.pone.0192420.

