## Supplementary material for "Extracellular Vesicles for Alzheimer’s Disease and Dementia Diagnosis": Table S1

**­­Table S1.** Complete search strategy using PUBMED and EMBASE. The search was performed from date of inception until Mar 22^nd^, 2024.

| PUBMED | ((exosome OR extracellular vesicle) AND (Alzheimer disease OR Alzheimer dementia)) |
| --- | --- |
| EMBASE | ('exosome'/exp OR 'exosome' OR 'extracellular vesicle'/exp OR 'extracellular vesicle') AND ('alzheimer disease'/exp OR 'alzheimer disease' OR 'alzheimer dementia') |

**­**

**­­**
